# Prostate cancer androgen receptor activity dictates efficacy of Bipolar Androgen Therapy

**DOI:** 10.1101/2022.04.26.22274275

**Authors:** Laura A Sena, Rajendra Kumar, David E Sanin, Elizabeth A Thompson, D Marc Rosen, Susan L Dalrymple, Lizamma Antony, Yuhan Yang, Carolina Gomes-Alexandre, Jessica L Hicks, Tracy Jones, Kiara A. Bowers, Jillian N Eskra, Jennifer Meyers, Anuj Gupta, Alyza Skaist, Srinivasan Yegnasubramanian, Jun Luo, W Nathaniel Brennen, Sushant K Kachhap, Emmanuel S Antonarakis, Angelo M De Marzo, John T Isaacs, Mark C Markowski, Samuel R Denmeade

## Abstract

Testosterone is the canonical growth factor of prostate cancer but can paradoxically suppress its growth when present at supraphysiological levels. We have previously demonstrated that the cyclical administration of supraphysiological androgen (SPA), entitled Bipolar Androgen Therapy (BAT), can result in tumor regression and clinical benefit for patients with castration-resistant prostate cancer [1–5]. However, predictors and mechanisms of response and resistance have been ill-defined. Here we show that growth inhibition of prostate cancer models by SPA requires high androgen receptor (AR) abundance and activity and is driven in part by downregulation of MYC. Using matched sequential patient biopsies, we show that high pre-treatment AR activity predicts downregulation of MYC, clinical response, and prolonged progression-free and overall survival for patients on BAT. BAT induced strong downregulation of AR in all patients, which is shown to be a primary mechanism of acquired resistance to SPA. Acquired resistance can be overcome by alternating SPA with the AR inhibitor enzalutamide, which induces adaptive upregulation of AR and re-sensitizes prostate cancer to SPA. This work identifies a predictive biomarker of response to BAT and supports a new treatment paradigm for prostate cancer involving alternating between AR inhibition and activation.

## Main

Signaling through the androgen receptor (AR) is the primary oncogenic driver of advanced prostate adenocarcinoma. Inhibition of AR signaling by androgen deprivation and AR inhibitors produces significant therapeutic and palliative benefit and constitutes the cornerstone of treatment of advanced disease. Yet this therapeutic strategy is not curative, and patients eventually progress with lethal castration-resistant prostate cancer (CRPC). Studies dating back 30 years indicate that supraphysiological androgen (SPA) can paradoxically suppress the growth of some human CRPC cell line and xenograft models [6,7]. These studies led us to test SPA as a treatment for men with CRPC. Our approach has been to pulse intramuscular testosterone every 28 days concurrent with ongoing luteinizing hormone-releasing hormone analogue administration to result in oscillation of serum testosterone from supraphysiological to near-castrate levels. Given this oscillation of testosterone between polar extremes, we termed this treatment Bipolar Androgen Therapy (BAT) [8]. We have previously described that BAT can produce clinical benefit and tumor regression for 20-30% of patients with CRPC [1–5], however predictors and mechanisms of response and resistance have been ill-defined.

To better understand growth-inhibitory mechanisms of SPA, we first studied human prostate cancer cell lines with varying sensitivity to SPA. SPA can be provided to cells as R1881, a potent synthetic androgen that is not metabolized *in vitro*, at a dose of 10nM, which is approximately 20-fold higher than the level of free testosterone in eugonadal adult men. LNCaP and VCaP cell lines are growth-inhibited by SPA, while LAPC4 and 22Rv1 cell lines exhibit primary resistance to SPA (Fig 1a) [1,9,10]. Among these cell lines, we observed that pre-treatment AR abundance and activity (as assessed by expression of the AR target PSA) was higher in SPA-sensitive cell lines than SPA-resistant cell lines (Fig 1b). High baseline AR abundance and activity was required for growth inhibition by SPA, as inducible shRNA-mediated knock-down of AR in LNCaP cells (Fig 1c) resulted in resistance to SPA (Fig 1d). Moreover, we found that high baseline AR abundance can also be sufficient to confer sensitivity to SPA, as overexpression of AR in LAPC4 cells (Fig 1e) resulted in growth inhibition by SPA (Fig 1f). This indicates that AR abundance and activity is a major determinant of prostate cancer response to SPA *in vitro*.

**Figure 1.**
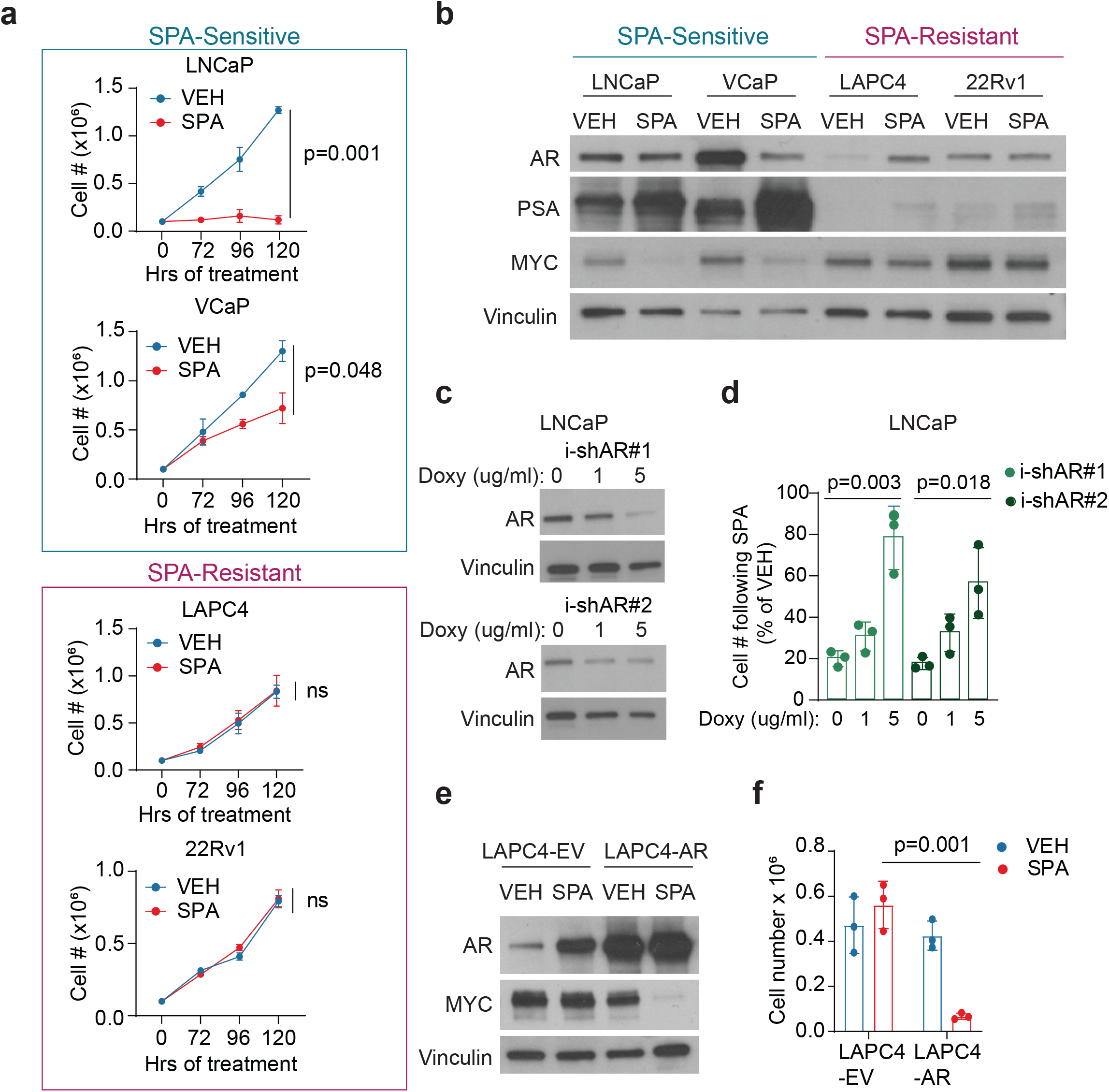
High pre-treatment AR activity is required and can be sufficient for growth inhibition by SPA. a, LNCaP, VCaP, LAPC4, and 22Rv1 viable cell number following treatment with VEH or SPA. p value by unpaired two-tailed t-test comparing final cell counts (n = 2 independent experiments). b, AR, PSA, and MYC protein expression by western blot of cell lines treated with VEH or SPA for 72 hours. Representative blot of n = 3 independent experiments. c, AR protein expression by western blot of LNCaP expressing inducible shRNA against AR pretreated with indicated concentration of doxycycline for 72 hours then VEH or SPA for 96 hours. Representative blot of n = 2 experiments. d, Viable cell count of LNCaP-shAR pretreated with indicated concentration of doxycycline for 72 hours then VEH or SPA for 96 hours. p value by unpaired two-tailed t-test comparing final cell counts (n = 3 independent experiments). e, AR and MYC protein expression by western blot of LAPC4-empty vector (LAPC4-EV) and LAPC4-AR cell lines treated with VEH or SPA for 7 days. Representative blot of n = 3 independent experiments. f, Viable cell count of LAPC4-EV and LAPC4-AR cell lines treated with VEH or SPA for 7 days (n = 3 independent experiments). VEH, vehicle control EtOH 0.01%. SPA, R1881 10 nM. i-shAR, inducible-short hairpin RNA against AR. For western blots (b, c, e), vinculin was used as a loading control.

AR activation has previously been shown to downregulate the growth-promoting factor c-MYC (hereafter called MYC) in normal prostate epithelial cells [11–13] and preclinical models of CRPC [14–17]. MYC is frequently upregulated in prostate cancer and is a critical driver of growth and proliferation [18,19]. We observed that SPA downregulates MYC, but only in cells lines with high AR abundance and activity (SPA-sensitive) and not in SPA-resistant cell lines (Fig 1b and Extended Data Fig 1a). High pre-treatment AR abundance was required for downregulation of MYC by SPA as inducible shRNA-mediated knock-down of AR in LNCaP cells disabled MYC downregulation by SPA (Extended Data Fig 1b). Moreover, high pre-treatment AR abundance was sufficient to induce downregulation of MYC by SPA given that AR overexpression resulted in MYC downregulation in LAPC4 cells (Fig 1e). Constitutive expression of MYC partially rescued growth inhibition of LNCaP and LAPC4-AR cells treated with SPA (Extended Data Fig 1c-e). These data suggest that SPA inhibits growth of CRPC with high AR abundance in part through downregulation of MYC. We anticipate that MYC-independent maladaptive effects of SPA may also contribute to growth inhibition, including AR inhibition of DNA relicensing during mitosis [20], AR-mediated DNA damage [21,22], and induction of ferroptosis and immunogenic cell death [10].

To assess molecular mechanisms of BAT in patients with mCRPC, we evaluated clinical samples from a prospective clinical trial (NCT03554317) that included on-study sequential paired biopsies of soft tissues metastases before (“pre-BAT”) and after 3 cycles of BAT (“on-BAT”) (Fig 2a). Twenty-four patients had tumor samples collected at both time points that were adequate for analysis by immunohistochemistry (IHC), and 15 of these patients had paired samples adequate for RNA sequencing analysis. Ten of 24 patients were considered to be responders, based on the presence of a decline in the serum PSA by at least 50% or tumor volume by at least 30% on cycle 4 day 1 of BAT. In pre-BAT samples, responders exhibited a non-significant trend toward higher total AR protein abundance by quantitative image analysis than non-responders (Fig 2b), which directly correlated with AR protein abundance in cytoplasm and nucleus, as well as *AR* mRNA abundance (Extended data Fig 2a-c). Prior to BAT, the nuclear-to-cytoplasmic ratio of AR was greater than 1 in all patients (Extended Data Fig 2d), indicating that the majority of AR resides in the nucleus in advanced mCRPC, which was not significantly different between responders and non-responders (Extended Data Fig 2d). To examine whether there was greater variation in pre-treatment AR activity between responders and nonresponders, we generated an AR activity score using Mann-Whitney ranking of expression of 10 canonical AR target genes (ARA_MW_ score) (Extended Data Fig 3a). Notably, responders had significantly higher pre-BAT ARA_MW_ scores than non-responders (p=0.011) (Fig 2c). The ARA_MW_ score was not driven by expression of one dominant gene (Extended Data Fig 3b), the included gene transcripts did not exhibit significant co-linearity (Extended Data Fig 3c), and the score did not correlate with AR protein abundance (Extended Data Fig 3d), indicating each included gene contributed unique data to the score, which overall was distinct from AR abundance. This demonstrates that AR activity is controlled by factors beyond protein abundance in advanced CRPC, which likely includes activating gene mutations in AR and activity and abundance of AR co-factors and regulators [23]. Stratifying patients by a cut-off ARA_MW_ score of 0.6 (selected due to its ability to stratify two distinct clusters of patients (Fig 2c) with distinct outcomes), we observed that patients with high (>0.6) ARA_MW_ scores exhibited greater PSA responses (p=0.010) (Fig 2d) and longer radiographic progression-free and overall survival (p=0.058 and p=0.002, respectively) (Fig 2e-f) on BAT. This indicates that pre-treatment AR activity is a major determinant of CRPC response to BAT, and the ARA_MW_ score may constitute a valuable predictive biomarker for this therapy.

**Figure 2.**
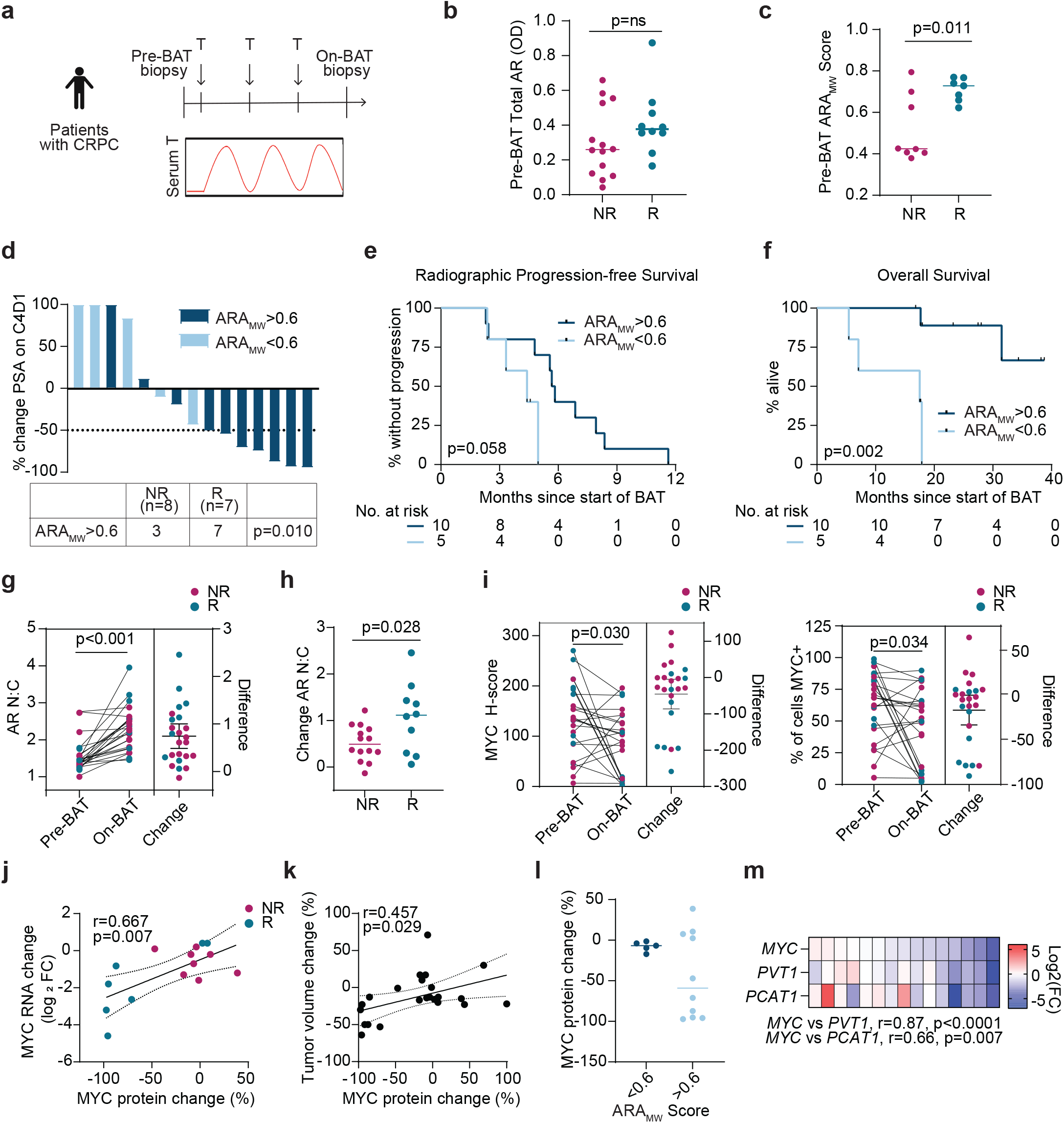
High pre-treatment AR activity is required for downregulation of MYC by BAT and predicts clinical benefit from BAT. a, Clinical trial design. CRPC, castration-resistant prostate cancer. BAT, Bipolar Androgen Therapy. T, testosterone. b, Pre-BAT total AR optical density (OD) by image analysis among non-responders (NR) and responders (R) with median indicated by line (n=24). Responders are those with a PSA_50_ response or objective response on C4D1. c, Pre-BAT ARA_MW_ score among NR and R with median indicated by line (n = 15). p value by unpaired two-tailed t-test. d, Percent change in PSA at C4D1 color-coded by ARA_MW_ score. PSA_50_ response indicated by dashed line. p value by Chi-squared comparison of proportions. e, Radiographic progression-free survival on BAT stratified by ARA_MW_ score. p value by log-rank. f, Overall survival on BAT stratified by ARA_MW_ score. p value by log-rank. g, AR nuclear-to-cytoplasmic ratio (AR N:C) in paired tumor biopsies (n=24 paired biopsies). p value by paired two-tailed t-test. Change with mean with 95% CI. h, Change in AR N:C among NR and R with median indicated by line (n=24). p value by unpaired two-tailed t test. i, MYC H-score and percentage of cells positive for MYC in paired tumor biopsies (n=24 paired biopsies). p value by paired two-tailed t-test. Change with mean with 95% CI. j, Correlation of *MYC* RNA change from C1D1 to C4D1 with MYC protein change from C1D1 to C4D1 (n = 15 paired biopsies). r and p values by Pearson’s correlation calculation. k, Correlation of percent change in tumor volume by RECIST measurement from C1D1 to C4D1 with MYC protein change from C1D1 to C4D1 (n = 23; 1 patient excluded for lack of measurable disease). l, Percent change in MYC protein expression stratified by ARA score (n = 15). m, Expression of genes within the 8q24 topologically associated domain (TAD) in patient tumor biopsy samples expressed as log_2_(fold change) from baseline pre-BAT (n = 15). r and p values by Pearson’s correlation calculation.

Strengths of the ARA_MW_ score are that it does not require a reference expression vector and is independent of differences in sequencing depth and processing. Therefore we applied ARA_MW_ scoring to an independent cohort of 266 patients with mCRPC who were not exposed to BAT (SU2C/PCF cohort) [24]. Among these patients, the prevalence of the biomarker (score >0.6) was 36.5% (Extended Data Fig 4a). If the biomarker is required for a PSA_50_ response to BAT and the PSA_50_ response rate among biomarker-positive patients is 70% (Fig 2d), then the predicted PSA response rate to BAT among patients with mCRPC is 25.6% (i.e., 36.5% x 70%). This is consistent with the measured PSA_50_ response rate to BAT of 24.3% among 173 patients with mCRPC across 2 clinical trials [3–5] (Extended Data Fig 4b), and provides plausibility that the ARA_MW_ score can predict response to BAT. Analysis of the SU2C/PCF cohort also indicated that the biomarker is not prognostic (i.e., it does not predict favorable outcomes independent of BAT treatment) (Extended Data Fig 5a) and is not clearly associated with particular genomic alterations or other patient factors (Extended Data Fig 5b-d).

We next assessed molecular changes induced by BAT in the paired sequential tumor biopsies. BAT increased the AR nuclear-to-cytoplasmic ratio in all patients (Fig 2g and Extended Data Fig 6a), but notably to a greater degree in responders (Fig 2h). This may suggest that nuclear recruitment/retention and/or cytoplasmic clearance of AR is important for a clinical response to BAT. We also examined MYC expression. Most patients had high expression of MYC protein in the pre-BAT tumor sample (Fig 2i). BAT decreased the median MYC expression, with a subset of patients having a near-complete ablation of MYC expression (Fig 2i and Extended Data Fig 6b). The change in MYC protein expression directly correlated with change in *MYC* mRNA expression (Fig 2j), suggesting that BAT suppressed MYC at the level of transcription and/or mRNA stability. BAT also decreased the median expression of the proliferation marker Ki-67 (Extended Data Fig 6c-d). The change in MYC protein expression directly correlated with the change in Ki-67 expression (Extended Data Fig 6d) and the change in tumor volume on CT scan (Fig 2k). Notably, only patients with pre-BAT ARA_MW_ scores greater than 0.6 exhibited significant decrease in MYC protein expression (Fig 2l), further supporting the concept that high pre-BAT AR activity is required for downregulation of MYC and tumor regression by SPA. The mechanism by which AR activation suppresses MYC in CRPC was recently suggested to occur through AR-mediated sequestration of cofactors and decreased activity of distal super-enhancers (SE) near *PCAT1* that regulate the *MYC* promoter, as well as those of neighboring transcripts embedded in the topologically associated domain (TAD) on 8q24 [17]. We noted that transcripts of genes located within the 8q24 TAD, *PCAT1* and *PVT1*, had similar change in expression as *MYC* on BAT (r=0.87, p<0.0001, and r=0.66, p=0.007, respectively) (Fig 2m), which supports an idea that BAT reduces *MYC* mRNA expression via disruption of distal SE activity.

Clinically we have observed that most patients with CRPC who initially respond to BAT acquire secondary resistance after approximately 6-12 months of therapy [5]. Similarly, the SPA-sensitive cell line LNCaP, which was initially cell cycle-arrested in G0-G1 after 5 days of SPA, re-entered the cell cycle following 12-19 days of continuous SPA exposure (Fig 3a). These cells were verified to have acquired resistance to SPA, as retreatment with increasing doses of R1881 resulted in no change to clonogenic survival (Fig 3b). The transcriptional and chromatin accessibility profiles of LNCaP with acquired resistance to SPA (LNCaP-SPAR) appeared most similar to VEH-treated cells (Extended Data Fig 7a-b), suggesting these cells revert to a pretreatment phenotype. Resistance did not appear to be driven by complete failure of SPA to activate AR, as Hallmark Androgen Response genes remained induced in both SPA-sensitive and resistant cells (Extended Data Fig 7c). Instead, development of resistance to SPA was associated with decreased expression of AR mRNA and protein, decreased AR activity assessed by decreased *KLK3* (encodes PSA) and PSA expression, and loss of suppression of *MYC* (Fig 3c-f). The AR promoter had reduced accessibility as early as 5 days of SPA, which persisted at 26 days (Fig 3g), consistent with prior reports indicating that ligand-bound AR exhibits negative autoregulation at the level of *AR* gene transcription [25]. MYC target gene sets were globally reactivated following development of resistance to SPA (Extended Data Fig 7d), and *MYC* re-expression was associated with re-expression of *PCAT1* and *PVT1* (Extended Data Fig 8a) and reorganization of 8q24 SE accessibility (Extended Data Fig 8b). To determine whether downregulation of AR was driving development of acquired resistance, we constitutively expressed full-length AR in LNCaP cells (Fig 3h). LNCaP-AR cells exhibited enhanced suppression of MYC by SPA (Fig 3h), followed by extensive vacuolization (Extended Data Fig 9) and cell death, not resistance (Fig 3i). This indicates that downregulation of AR is a mechanism of acquired resistance to SPA *in vitro*.

**Figure 3.**
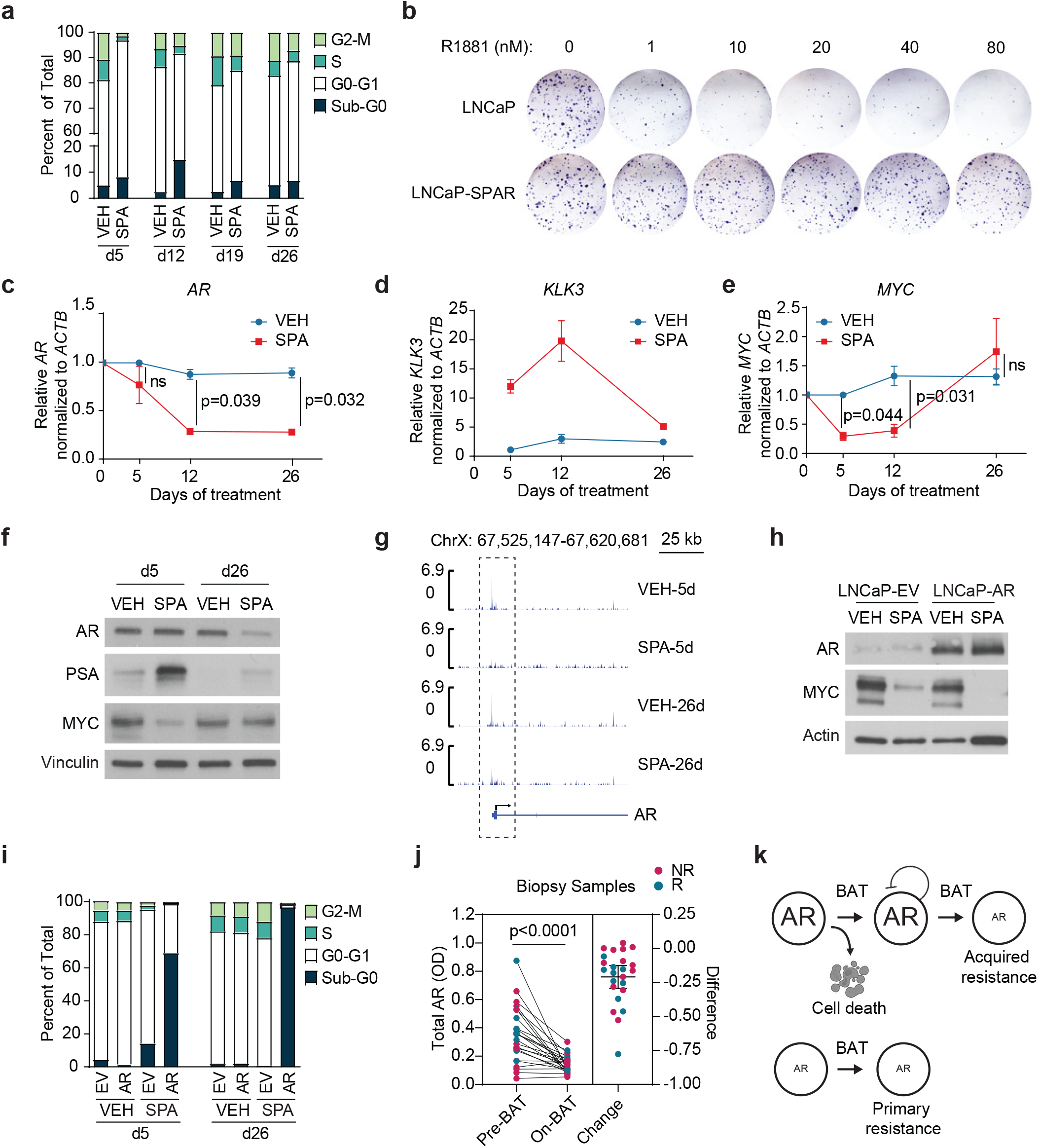
Downregulation of AR can drive acquired resistance to SPA. a, Cell cycle analysis by propidium iodide staining of LNCaP cells treated with VEH or SPA. Average values of n = 2 independent experiments. b, Clonogenic survival of LNCaP cells treated for 26 days with VEH or SPA, followed by 7 days rest without treatment, followed by treatment with dose of R1881 as indicated. Representative photograph of n = 2 independent experiments. c-e, AR, KLK3, MYC mRNA expression by qPCR of LNCaP cells treated with VEH or SPA (n = 3 independent experiments). Ct-values were normalized to ACTB for each sample, then to VEH x 5 days, and expressed as median ± s.e.m with p values by unpaired two-tailed t-test. f, AR, PSA, and MYC protein expression by western blot of LNCaP cells treated with VEH or SPA for 5 or 26 days. Representative blot of n = 3 independent experiments. g, Chromatin accessibility by ATAC-seq of the AR promoter (dotted box) of LNCaP cells treated with VEH or SPA for 5 or 26 days (performed in duplicate). h, AR and MYC protein expression by western blot of LNCAP-EV and LNCAP-AR cells treated with VEH or SPA for 72 hours. Representative blot of n = 3 independent experiments. i, Cell cycle analysis by propidium iodide staining of LNCaP-EV and LNCaP-AR cells treated with VEH or SPA. Average values of n = 3 independent experiments. j, Total AR optical density (OD) by image analysis in paired tumor biopsies (n=24 paired biopsies) color coded by non-responder (NR) or responder (R). p value by paired two-tailed t-test. Change with mean with 95% CI. k, Schematic model of primary and acquired resistance to BAT. VEH, vehicle control, EtOH 0.01%. SPA, R1881 10 nM, LNCaP-SPAR, LNCaP with acquired resistance to SPA. For western blots, vinculin (f) or actin (h) was used as loading controls.

Returning to the patient samples, we saw that BAT induced strong downregulation of AR mRNA and protein in all patients (Fig 3j and Extended Data Fig 10a), with higher pre-BAT AR predicting greater decrease by BAT (Extended Data Fig 10b). This might be explained by a threshold effect, meaning that BAT decreases AR to a threshold minimum level below which AR is not further suppressed by BAT. These data demonstrate that adaptive downregulation of AR occurs in patients and might lead to acquired resistance to BAT over time. This mechanism is consistent with an emerging conceptual idea that acquired resistance to cancer therapy is often driven by plastic (i.e. reversible) cellular alterations, rather than gene mutation [26]. Overall these data suggest that low AR expression and activity is a mechanism of primary and acquired resistance to BAT (Fig 3k).

We have previously reported that patients who have progressed on BAT appear to have enhanced clinical responses to subsequent AR inhibition [2–5]. For example, in the TRANSFORMER study, patients with mCRPC who had not received prior BAT exhibited a PSA_50_ response rate of 25% and median response duration of 3.8 months to the AR inhibitor enzalutamide, while patients who had progressed on BAT exhibited a PSA_50_ response rate of 78% and median duration of response of 10.9 months to enzalutamide [5]. Similarly, LNCaP-SPAR cells were more growth-inhibited by enzalutamide compared to parental LNCaP (Fig 4a-b). This may be due to reduction of AR abundance by prior treatment with SPA (Fig 3f) that can sensitize cells to AR inhibition. Notably, enzalutamide treatment resulted in adaptive upregulation of AR in both cell lines (Fig 4c) and enhanced downregulation of MYC (Fig 4d) and growth inhibition by subsequent treatment with SPA (Fig 4e). This indicates that response to SPA can be restored after development of acquired resistance through use of AR inhibitors like enzalutamide to induce adaptive upregulation of AR.

**Figure 4.**
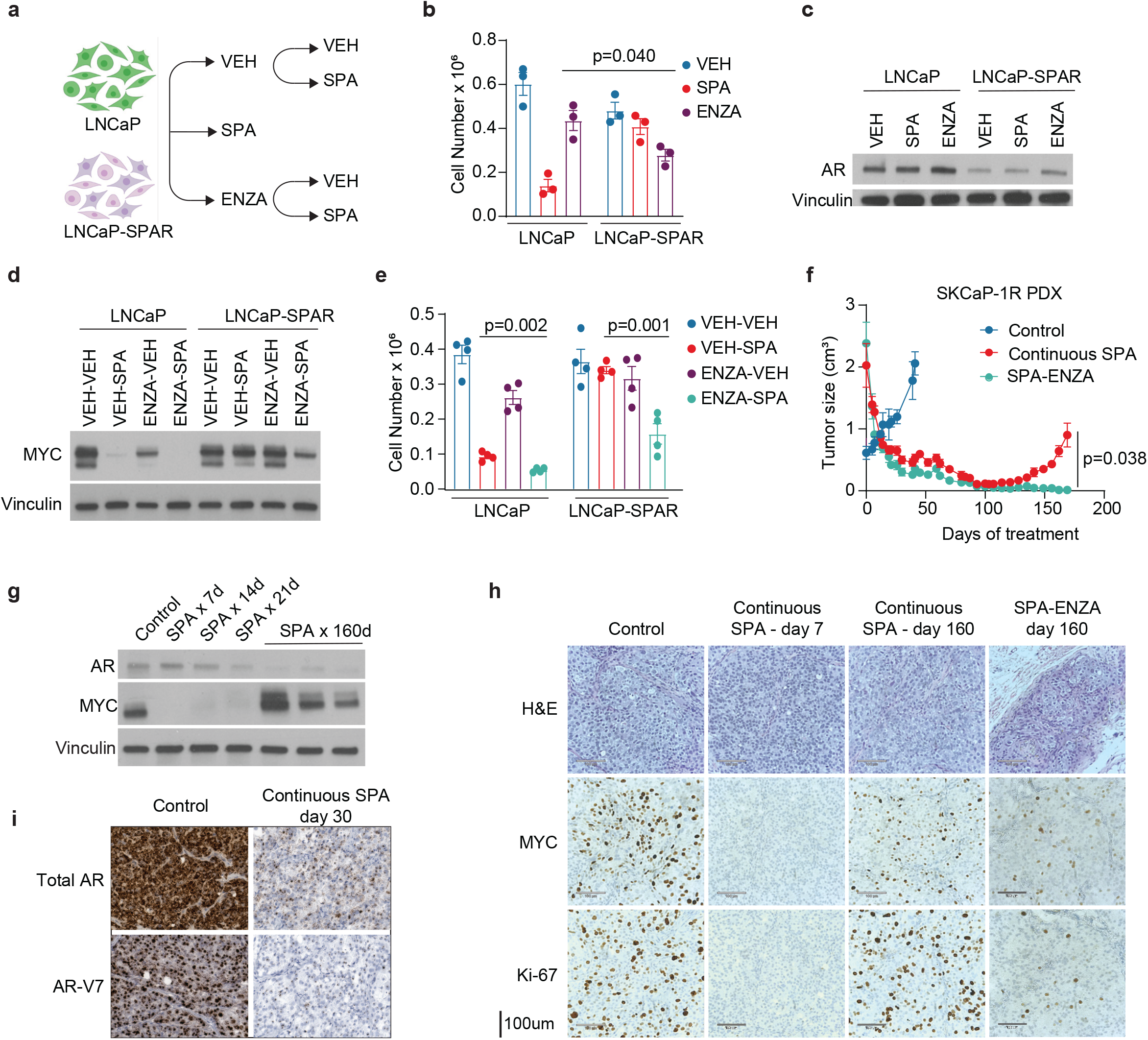
Acquired resistance to SPA can be overcome by alternating between AR activation and inhibition. a, Experimental design schematic. LNCaP-SPAR are LNCaP cells with acquired resistance to SPA. b, Viable cell number of LNCaP and LNCaP-SPAR cells following treatment with VEH, SPA, or ENZA for 5 days (n = 3 independent experiments). p value by unpaired two-tailed t-test. c, AR protein expression by western blot of cells treated as per a. Representative blot of n = 3 independent experiments. d, MYC protein expression by western blot of LNCaP and LNCaP-SPAR cells following treatment with VEH or ENZA for 5 days followed by VEH or SPA for 5 days. Representative blot of n = 3 independent experiments. e, Viable cell number of cells treated as per d (n = 4 independent experiments). p values by unpaired two-tailed t-test. f, Tumor size of SKCaP-1R patient-derived xenograft (PDX) following no treatment (Control; n = 4 mice), continuous testosterone (SPA; n = 4 mice), or SPA alternating with enzalutamide every 3 weeks (SPA-ENZA; n = 3 mice). p value by unpaired two-tailed t-test comparing final measurements. g, AR, and MYC protein expression by western blot of SKCaP-1R untreated (control) or treated with SPA. h, Hematoxylin and eosin (H&E) staining and immunohistochemistry for MYC and Ki-67 of SKCaP-1R following no treatment (Control), continuous SPA for 7 or 160 days, or SPA-ENZA for 160 days. Representative photograph of n = 3 mice per group. i, RNA in situ hybridization for AR and AR-V7 in tumors of SKCaP-1R untreated (control) or treated with SPA for 30 days. Representative photograph of n = 3 mice per group. For western blots (c, d, g) vinculin used as a loading control.

While BAT was originally designed to cycle testosterone levels to minimize adaptation to high or low levels of androgens, these data suggest that there may be clinical benefit to more extreme oscillation of AR activity by alternating the use of SPA with an AR inhibitor. To test this therapeutic strategy, we used a patient-derived xenograft (PDX) model derived from a metastasis of a patient with CRPC that was adapted to grow in a castrated mouse (SkCaP-1R) [27]. This PDX, which expresses high AR and the AR splice-variant AR-V7 and is resistant to the second generation androgen signaling inhibitors abiraterone and enzalutamide [27], initially regressed in response to SPA, but acquired resistance after about 5 months (Fig 4f). Regression was associated with an almost complete loss of MYC expression, while acquired resistance was associated with a decrease in AR and return of MYC expression (Fig 4g-h). SPA suppressed mRNA expression of *AR* and *ARV7* as early as 21-30 days (Fig 4g-i). Notably, when SPA was alternated every 21 days with enzalutamide (SPA-ENZA), this PDX did not acquire resistance after 160 days of observation (Fig 4f). Subcutaneous tissues from the animals that received SPA-ENZA for 160 days were analyzed histologically and nests of cancer cells were observed, but these cells lacked significant staining for the proliferation marker Ki-67 (Fig 4h). These data suggest that repeat cycling of SPA and AR inhibition may prevent the development of adaptive resistance and lead to more durable growth inhibition of prostate adenocarcinoma. To determine the benefit of this strategy for patients with CRPC, we have initiated a clinical trial in which patients are repeatedly switched from BAT to enzalutamide to BAT, entitled, “Sequential Testosterone and Enzalutamide Prevents Unfavorable Progression (STEP-UP)” (NCT04363164). By shifting the treatment strategy to anticipate resistance, this dynamic protocol is an attempt to use game theory [28] to minimize resistance and maximize physician control of prostate cancer growth.

Our results indicate that high AR abundance and activity is required for growth inhibition of prostate cancer by SPA, which occurs in part through downregulation of MYC. We identify the ARA_MW_ score as a predictive biomarker of response to BAT, which should be validated in future clinical studies, but may be valuable for patient selection for BAT. Primary and acquired resistance to SPA can be driven by low abundance of AR, but acquired resistance can be overcome by treatment with an AR inhibitor, which induces upregulation of AR and increases susceptibility to SPA. This adaptive upregulation of AR by AR-axis inhibitors is well described to occur in patients and constitutes a mechanism of resistance to these agents [29,30]. We show here that BAT conversely downregulates AR, and this resulted in enhanced sensitivity to enzalutamide. As such, prostate cancer engages in classical endocrine negative feedback loops to titrate AR to abundance of ligand, and our results suggest this can be exploited therapeutically. While the status quo as it pertains to treatment of prostate cancer is persistent and potent AR inhibition, this work provides rationale to alternate between AR inhibition and activation with BAT to prolong the lives of patients with this disease.

## Methods

### Cell culture and reagents

LNCaP and VCaP cell lines were obtained from American Type Culture Collection (ATCC). LAPC4 and 22Rv1 cell lines were a gift from J Isaacs. LNCaP, LAPC4, and 22Rv1 were grown in RPMI 1640 (Gibco; 11835-055) supplemented with 10% fetal bovine serum (Corning), sodium lactate 1.6mM, sodium pyruvate 0.5mM, Lalanine 0.43mM, 1% pen-strep (Gibco). VCaP was grown in DMEM (ATCC 30-2002) supplemented with 10% fetal bovine serum (Corning) and 1% pen-strep (Gibco). Full-length AR cDNA was cloned into the BAMH1/Sal1 site of pLenti-CMV-GFP-Puro (Addgene; 17448, E Campeau and P Kaufman laboratories) and empty vector control was generated through excision of GFP of this plasmid. These vectors, along with pCDH-puro-cMyc vector (Addgene; 46970, J Wang laboratory), pCDH-EF1-FHC empty vector control (Addgene, 64874, R Wood laboratory), piSMARTvector-PGK-TurboGFP-TRE3G-shAR vectors (Horizon; V3IHSPGG_8216343 and V3IHSPGG_7292640) were transfected into 293T cells (ATCC) along with pMD2.G (Addgene; 12259) and psPAX2 (Addgene; 12260) packaging vectors using lipofectamine (Invitrogen) to produce AR-puro, cMyc-puro, control-puro, and Tet-On-shAR-puro lentivirus particles. Two days after transduction with indicated virus, vector-expressing cells were selected with puromycin 1ug/ml for 72 hours. To constitutively express MYC in LAPC4-AR cells, LAPC4-AR cells were transfected with pCDNA3-HA-HA-humanCMYC vector (Addgene; 74164, M Roussel laboratory) using lipofectamine (Invitrogen) then selected for stable expression with G418 (Sigma) for 7 days. Cells were maintained at 37 °C in 5% CO2. They regularly tested negative for mycoplasma contamination using MycSensor PCR Assay kit (Agilent Technologies). R1881 was obtained from Sigma, enzalutamide from Selleck Chem, and testosterone cypionate from Steraloids.

Proliferation, cell viability, clonogenic survival, and cell cycle analyses of cell lines. For proliferation and cell viability assays, cells were plated in triplicate on 6-well (0.1 × 10^6^ cells/well) or 12-well (0.05 × 10^6^ cells/well) plates and incubated with R1881, enzalutamide, or vehicle control for indicated duration. Cells were counted using a hemocytometer with viability assessed by trypan blue exclusion. For clonogenic survival assessment, cells were plated on 6-well plates at low density (1500 cells/well) in 1 ml fresh media and 250 ul conditioned media obtained from a confluent flask of the parental cell line. Colonies were stained with crystal violet (Sigma; 0.5% in 20% methanol) after 10-20 days. For cell cycle analysis, cell pellets were resuspended in cold 70% ethanol at least overnight, then subsequently washed with PBS, and resuspended in 50 ug/ml propidium iodide (Sigma) and 100 ug/ml RNAse (Sigma) and run on the BD FACSCelesta flow cytometer with analysis using FlowJo software 10.4.2.

### Western blot analyses

Cells or tissues were lysed with 1x denaturing lysis buffer (Cell Signaling technology) containing protease and phosphatase inhibitors (Roche). Protein concentration was determined using the Pierce BCA Protein Assay kit (Thermo Scientific), and 5-20 ug of lysate was resolved on a SDS-PAGE gel and transferred to a nitrocellulose membrane. Membranes were blocked with 5% milk for 1 hour, then incubated with primary antibody overnight. Primary antibodies used were: anti-cMYC (Abcam, clone Y69, 1:1000 dilution), anti-AR (Santa Cruz, sc7305, 1:1000 dilution), anti-PSA (Dako, A0562, 1:1000 dilution), anti-vinculin (Millipore, clone V284, 1:2000 dilution). Anti-rabbit IgG, HRP-linked Antibody (CST, 7074S, 1:5000 dilution) and Anti-mouse IgG, HRP-linked Antibody (CST, 7076S, 1:5000 dilution) were used as secondary antibodies.

### Quantitative real-time polymerase chain reactions

RNA was extracted using the RNeasy kit (Qiagen) and cDNA generated using the high-capacity cDNA reverse transcription kit (ThermoFisher). RT-PCR were performed in triplicate using 500ug cDNA, 10ul TaqMan Gene Expression Master Mix (ThermoFisher), and 1ul 20x TaqMan Gene Expression Assay probe mix for *MYC* (Hs00153408_m1), *AR* (Hs00907244_m1), *KLK3* (Hs02576345_m1), and *ACTB* (Hs01060665_g1) (ThermoFisher) on an ABI7500 RT-PCR System (ThermoFisher). Relative gene expression was determined by delta-delta CT.

### Clinical trial design and procedures

The COMbination of Bipolar Androgen Therapy and Nivolumab (COMBAT-CRPC; NCT03554317) clinical trial was a single-arm, multicenter, open-label phase II study of BAT in combination with the anti-PD1 agent nivolumab for patients with metastatic CRPC that had progressed on at least one novel androgen receptor-target therapy. This study was approved by the Institutional Review Board at Johns Hopkins, and all accrued patients provided written informed consent. The inclusion and exclusion criteria and pre-specified study end points were previously described [31]. Patients were required to have soft tissue metastases amenable to biopsy to participate. Patients were treated with 3 cycles of BAT (testosterone cypionate 400 mg intramuscular every 28 days) followed by concurrent BAT and nivolumab 480 mg intravenously every 28 days until progression. Paired core-needle tumor biopsies were performed prior to treatment and after 3 cycles of BAT monotherapy. Response was assessed with PSA at each cycle, and CT chest, abdomen, and pelvis and technetium-99 bone scan every 3 cycles.

### Chromogenic Immunohistochemistry (IHC) for AR

Tissue sections were baked on a 60°C hotplate for 10 min. and then deparaffinized in xylenes. Tissues were rehydrated in graded alcohol solutions and distilled water (diH_2_O). Slides were then rinsed in a solution of diH_2_O with 0.1% Tween and placed in a citrate bucket wash for 1 min. Tissue sections were steamed in Target Retrieval Solution (Dako; S1699, Santa Clara, CA), washed in Tris Buffered Saline with Tween (TBST; Sigma), and subjected to a Dual Endogenous Enzyme Block (Dako; S2003). The tissues were incubated with the primary antibody (conditions varied for each antibody) and, rinsed with TBST, and incubated with the secondary antibody, PowerVision Poly-HRP anti-Rabbit IgG (Leica, Deer Park, IL), for 30 min. at RT. Tissues were rinsed with TBST and then aminoethyl carbazole (Immpact AEC; Vector Labs; SK-4205, Burlingame, CA) was applied for 20 min. at RT. Slides were washed with TBST and counterstained with hematoxylin (Mayers; Dako; S3309; diluted 1:4). Slides were then coverslipped with VectaMount AQ Aqueous Mounting Medium (Vector Laboratories; H-5501) and scanned on a Ventana DP200 Digital Whole Slide Scanner (Roche Diagnostics, Rotkreuz, Switzerland) using a 40X microscope objective. Coverslips were removed with diH_2_O and AEC was dissolved with graded levels of alcohol. Slides were then steamed and incubated with an SDS-2ME solution as previously described [32]. Slides were placed in TBST for 5 min. and then steamed, after which the dual endogenous block was reapplied and the tissue was incubated with the next antibody. This process was repeated until each tissue had been iteratively stained with the following antibodies:

| Staining Iteration | Antibody (Source) | Species | Clone | Dilution | Incubation Conditions |
| --- | --- | --- | --- | --- | --- |
| 1 | AR (Cell Signaling Technology) | Rabbit Monoclonal | D6F1I | 1:10,000 | overnight, 4°C |
| 2 | AR (Cell Signaling Technology) | Rabbit Monoclonal | D6F1I | 1:5,000 | overnight, 4°C |
| 3 | CK8 (Abcam) | Rabbit Monoclonal | EP1628Y | 1:3,000 | 45 min., RT |

As a control, after primary antibody removal, for each of the antibody staining rounds in a number of experiments we performed staining by leaving out the primary antibody and performing a round of staining with the secondary antibody only. These experiments showed a complete absence of signals indicating a complete antibody removal.

### Quantitative Image Analysis for AR Protein

Image analysis was performed using the MultiPlex module on HALO 3.2 (Indica Labs). Whole slide scans of each staining round for AR and CK8 were imported into HALO and registered with one another. A patientspecific Random Forest classifier was manually trained to classify all regions of each CK8-stained tissue into one of three categories: tumor, non-neoplastic tissue, and background. Annotations created from this classifier were then mapped onto the scans of the AR-stained tissues, using HALO’s “Classify Registered” feature. In the event that the scan of the CK8-stained tissue could not be successfully used for tumor classification (low CK8 expression, degraded tissue or imprecise scan registration), a patient-specific tumor classifier was trained on the scan of the AR-stained tissue. If the AR signals at a dilution of 1:10,000 was too weak for classifier training, scans of the tissues stained for AR at 1:5,000 were utilized to train the classifier. AR positivity thresholds were determined using the real-time tuning window. The MultiPlex analysis was restricted to tumor positive regions (using the CK8-trained classifier) and provided several measurements for each tumor-positive cell within each AR-stained tissue, including nuclear AR optical density (OD), cytoplasmic AR OD, nuclear area (µm^2^), cytoplasmic area (µm^2^), and cell area (µm^2^). Whole cell average AR ODs were determined by calculating an integrated cell OD for each cell and dividing that value by each cell’s respective area.

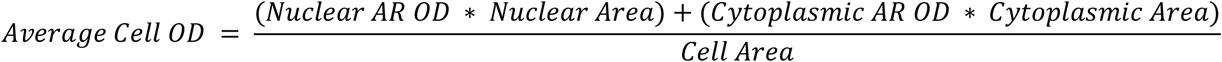

### Immunohistochemistry (IHC) for MYC and Ki67 Proteins

IHC on patient biopsy samples for MYC and Ki67 was performed as previously described [18,33]. IHC on SKCaP PDX tissue was performed by the Sidney Kimmel Comprehensive Cancer Center (SKCCC) IHC Core facility. Primary antibodies were anti-MYC (Epitomics, Clone Y69, 1:200) and anti-Ki67 (AbCam, Clone SP6, 1:200), and tissues were stained with the Ventana Discovery Ultra Autostainer, using the Discovery anti-HQ HRP kit.

### Quantitative Image Analysis for MYC and Ki67 Proteins

For quantitative image analysis, whole IHC stained biopsy slides were scanned using a 40X objective on a Ventana DP200 slide scanner. Whole slide scans were imported into HALO 3.1 image analysis platform. The nuclear staining for both MYC and Ki67 were analyzed using the cytonuclear module v 1.6. Tumor regions were delineated by a pathologist (CG-A) using manual annotations, guided by adjacent H&E and androgen receptor stained slides. Positively stained (DAB staining above minimum intensity threshold) for MYC and Ki67 cells were binned into 3 intensity levels. We calculated the percentage of positively stained cells for both Ki67 and MYC proteins, as well as an H-SCORE for MYC using the intensity levels with the following formula (percentage of intensity level 3 cells x 3)+(percentage of intensity level 2 cells x 2)+ (percentage of intensity level 1 cells x 1) to give a total possible score of 300. Percentage positively stained cells were validated on a number of samples by manual counting.

### Frozen biopsy specimens for laser capture microdissection (LCM) for bulk RNAseq analysis

Frozen sections were cut onto PEN (polyethylene naphthalate) slides (Leica), stained with hematoxylin and LCM was performed at the SKCCC Core facility using a Leica LMC7000 laser capture microdissection system in the SKCCC Cell Imaging Core. Laser captured tissues from regions highly enriched for tumor cells were collected by manual annotations of regions of interest that outlined tumor nests. RNA was isolated using the ALLPREP RNA/DNA extraction protocol (Qiagen Cat. No. 802804). RNA amounts were measured using a Qubit fluorometer (Invitrogen) and quality were measured using an Agilent Bioanalyzer 2100 with the Pico Chips Kit.

### RNA sequencing

For RNA sequencing of patient biopsy samples, purified RNA was provided to the SKCCC Experimental and Computational Genomics Core to carry out their low-input RNA-seq workflow as described previously with some modifications [34]. Briefly, quality of total RNA was measured by the Agilent Bioanlayzer to determine RNA integrity (RIN). Samples with starting input between 100pg-100ng of total RNA and RIN > 7.0 were considered to have sufficient quality to proceed to construction of whole transcriptome sample-barcoded libraries using the Ovation RNA-Seq System V2 according to the manufacturer’s protocols (Nugen).

Quantification of the libraries was performed by qPCR or by the Agilent Bioanalyzer and equimolar concentrations of each library were pooled together, clustered and sequenced on an Illumina Novaseq 6000 platform, with paired end sequencing. The resulting reads were aligned the human reference genome build hg38 using RSEM [35] with the STAR aligner [36] to obtain posterior mean estimate transcripts per million (pmeTPM) as a measure of gene expression. pmeTPM values for each gene were normalized using upper quantile normalization across samples and log2 transformed. For RNA sequencing of cell lines, purified RNA was sent to Admera Health who performed library preparation with polyA selection, sequencing on the Illumina HiSeq 2500 with read length configuration of 2×150bp with 60 million reads per sample, and read mapping to GRCM38. Sequenced libraries were processed with deepTools [37], using STAR [36], for trimming and mapping, and featureCounts [38] to quantify mapped reads. Raw mapped reads were processed in R (Lucent Technologies) with DESeq2 [39] to generate normalized read counts to visualize and cluster as heatmaps using pheatmap [53] and determine differentially expressed genes with greater than 2-fold change and lower than 0.01 adjusted p value. Gene set enrichment analysis was performed using fGSEA [40].

### ARA_MW_ Score Calculation

We calculated the AR activity score using Mann-Whitney ranking (ARA_MW_) by adapting a previously reported approach [41]. First, genes were selected based on previous reports [42] and examined for co-linearity based on the pearson correlation coefficient and testing for its significance using the cor and cor.test functions in R, respectively. Having observed no correlation between most genes, we selected 10 genes (*KLK2, KLK3, FKBP5, STEAP1, STEAP2, PPAP2A, RAB3B, ACSL3, NKX3-1, TMPRSS2*) and applied the adapted gene scoring method to RNAseq results from COMBAT biopsies, and to polyA captured RNAseq results from 266 biopsies from participants in the SU2C/PCF study [24] retrieved from cBioPortal [43]. Briefly, we used the log transformed quantile normalized pmeTPM values and calculated a ranked list of genes for each biopsy to generate a *g x b* matrix *R* of ranked genes (with *g* genes and *b* biopsies), considering only the top 1500 highest ranked genes by setting *r*_*g,b*_ = *r*_*max*_ + 1, with *r*_*max*_ = 1500. The ARA_MW_ score was then calculated for the *n* selected genes as:

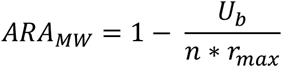

where *U* is the Mann-Whitney U statistic calculated as:

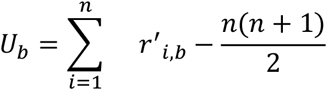

and *R*^′^ is a subset of *R* including only the *n* selected genes in the score. In this manner, the score was independent of sequencing depth, normalization strategy and dataset composition, and was primarily based on the relative abundance of selected genes within each studied sample.

Clinical data from 266 participants in the SU2C/PCF was retrieved from cBioPortal and participants classified based on ARA_MW_. Differences between classified patients for each clinical parameter was examined using a Wilcoxon ranked sum test.

### ATAC sequencing

Cells were provided to the Single Cell and Transcriptomics Core at Johns Hopkins University who performed nuclei isolation, library preparation, and sequencing with read length configuration of 2×75 with 50 million reads per sample. Briefly, 5×10^4^ cells were washed in PBS and then lysed in 10 mM Tris-HCl, pH 7.4,10 mM NaCl, 3 mM MgCl_2_ and 0.1% Igepal CA-630 (all SIGMA). Nuclei were then spun down and then resuspend in 25 μL TD (2x reaction buffer), 2.5 μL TDE1 (Nextera Tn5 Transposase) and 22.5 μL nuclease-free water, incubated for 30 min at 37°C. Purified DNA was PCR-amplified, then cleaned from adaptors. Sequenced libraries were trimmed with Trimmomatic [44], mapped using Bowtie2 [45], and processed with SAM tools [46] to retain open chromatin fragments of less than 150kbp. Coverage files were generated with deepTools [37]. Open chromatin and differentially regulated peaks were detected with MACS2 [47] with a p value < 1 × 10^−7^ and a q value of less than 0.1 and DiffBind [48] using a 2-fold change and 0.01 p value as significance thresholds. Bed files were analyzed with Bedtools [49], and visualized alongside coverage files on IGV [50]. Reference H3K27Ac CHIP-seq on human VCAP data was obtained from the ENCODE project [51].

### RNA in situ hybridization (RISH)

RISH was performed as previously described [52].

### Mouse models and tumor studies

Male NSG mice aged 8-12 weeks were obtained from the Sidney Kimmel Comprehensive Cancer Center (SKCCC) Animal Core Facility and surgically castrated. SKCaP patient-derived xenograft tissue was minced, mixed with Matrigel (BD Biosciences) and implanted subcutaneously on the flank. Testosterone was administered by implantation of a slow-release subcutaneous pellet in the opposite flank. Pellets were assembled using 25 mm sections of Silastic Laboratory tubing, filled with 30 mg testosterone cypionate (Steraloids INC A6960-000), sealed on both ends with Silastic Medical Adhesive Type A, then sterilized. Enzalutamide (Selleck Chem) was administered by oral gavage 10 mg/kg/day in 200 ul 1% carboxymethyl cellulose, 0.1% Tween-80, 5% DMSO. Tumors were measured twice weekly using microcalipers, and tumor volume was calculated using the following formula: 0.5236 x L x W x H. At study completion, mice were euthanized and tumors were extracted. Tumors were flash frozen for subsequent lysis for immunoblot analyses and formalin-fixed for subsequent immunohistochemistry analyses. All mice were housed in the Johns Hopkins animal facility and we have complied with all relevant ethical regulations in accordance with Johns Hopkins Institutional Animal Care and Use Committee.

## Data Availability

All data produced in the present study are available upon reasonable request to the authors

## Acknowledgements

We thank the members of the Sidney Kimmel Comprehensive Cancer Center’s Experimental and Computational Genomics Core, supported by NIH/NCI grant P30CA006973, for support in carrying out RNA-seq experiments and analysis of the paired biopsy specimens. We thank the members of the Johns Hopkins University Transcriptomics and Deep Sequencing Core for support in performing ATAC-seq experiments. This work was supported by Department of Defense early investigator awards to LAS and RK (W81XWH2010079 and PC210095, respectively), a JHU Clinician-Scientist award to LAS, a Prostate Cancer Foundation young investigator award to RK, an NIH/NCI SPORE prostate cancer grant (P50CA058236) to SRD, a Prostate Cancer Foundation challenge award to ESA and SKK, and W81XWH1910724 and 1R01CA243184-01A1 to SKK. We especially thank the patients and their caregivers who participated in the clinical trial described.

---

**Extended Data Figure 1.**
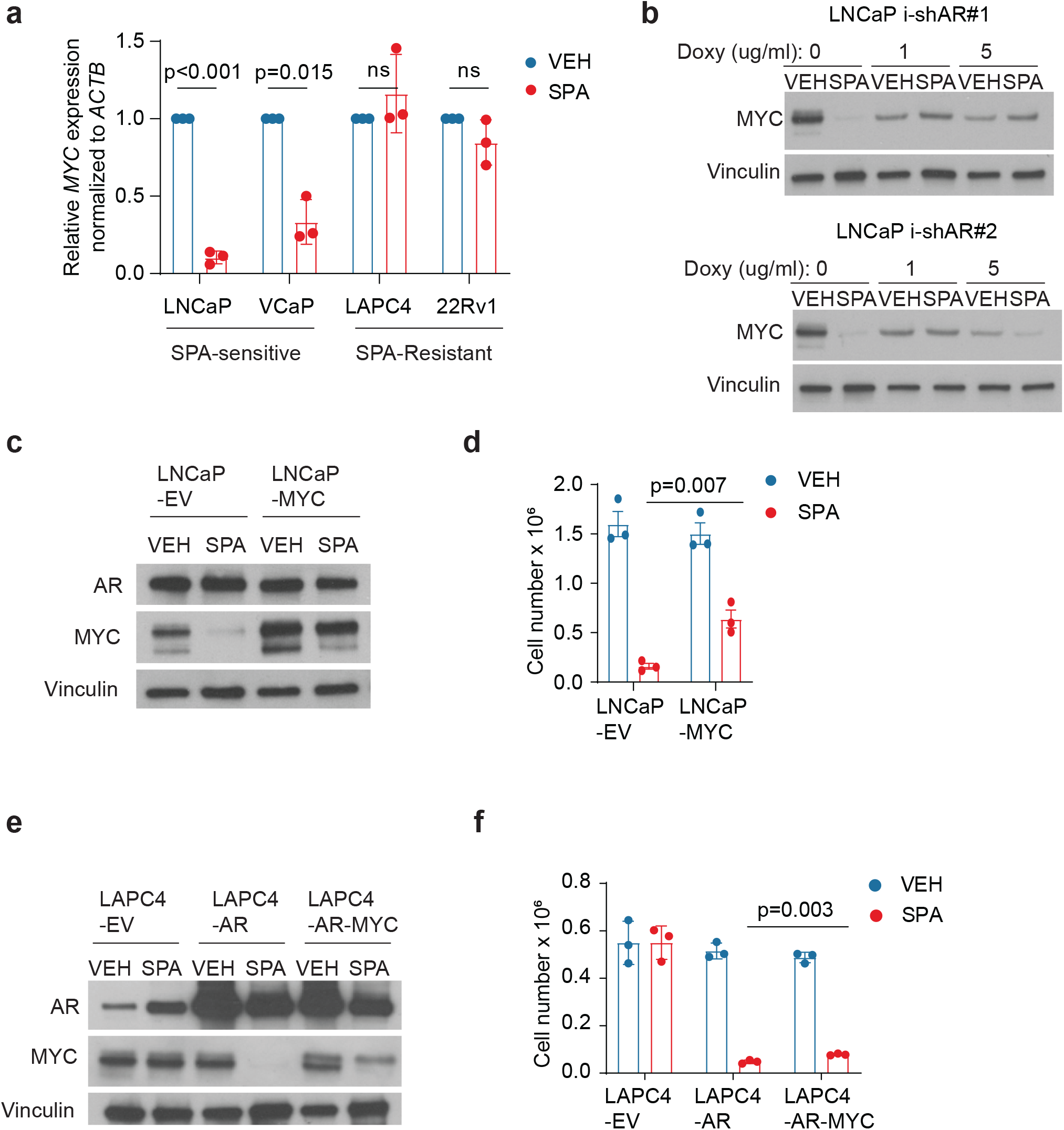
High pre-treatment AR is required for downregulation of MYC by SPA, which contributes to growth inhibition. a, MYC mRNA expression by quantitative PCR (qPCR) of prostate cancer cell lines treated with VEH or SPA for 72 hours (n = 3 independent experiments). Ct-value was first normalized to ACTB for each sample, then to VEH for each cell line, and expressed as median ± s.e.m with p values by unpaired two-tailed t-test with Welch correction for unequal variances. b, MYC protein expression by western blot of LNCaP expressing inducible shRNA against AR pretreated with indicated concentration of doxycycline for 72 hours then VEH or SPA for 96 hours. Representative blot of n = 2 experiments. c, AR and MYC protein expression by western blot of LNCaP-empty vector (LNCaP-EV) and LNCaP-MYC cell lines treated with VEH or SPA for 72 hours. Representative blot of n = 3 independent experiments. d, Viable cell count of LNCaP-EV and LNCaP-MYC cell lines treated with VEH or SPA for 7 days (n = 3 independent experiments). P value by unpaired two-tailed t-test. e, AR and MYC expression by western blot of LAPC4-EV, LAPC4-AR, and LAPC4-AR-MYC cell lines treated with VEH or SPA for 7 days. Representative blot of n = 3 independent experiments. f, Viable cell count of LAPC4-EV, LAPC4-AR, and LAPC4-AR-MYC cell lines treated with VEH or SPA for 7 days (n = 3 independent experiments). p value by unpaired two-tailed t-test. VEH, vehicle control, EtOH 0.01%. SPA, R1881 10 nM. For western blots (b, c and e), vinculin was used as a loading control.

**Extended Data Figure 2.**
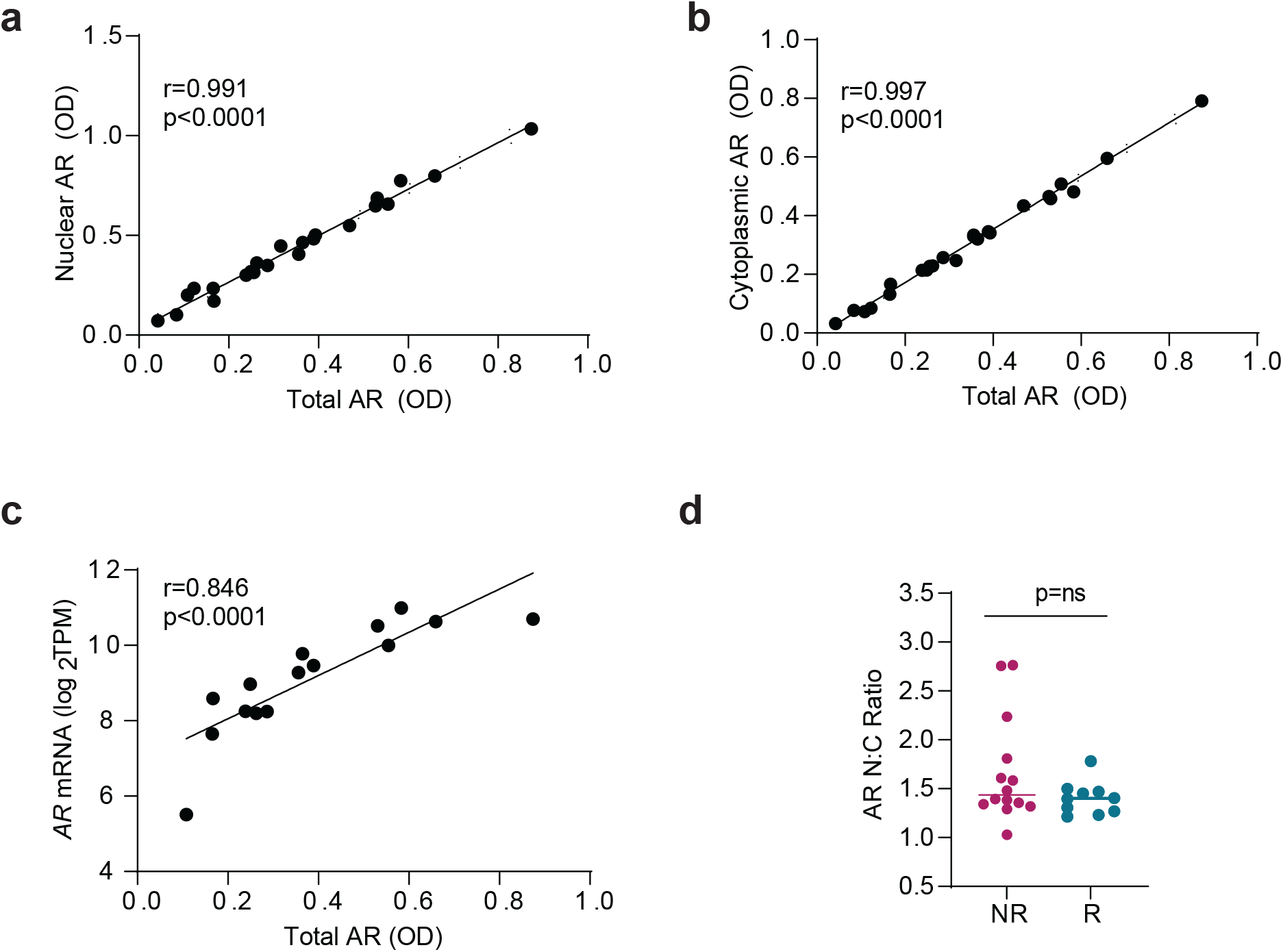
Total AR protein abundance correlates with nuclear AR, cytoplasmic AR, and *AR* mRNA. a, Correlation of nuclear AR with total AR within pre-BAT biopsies (n=24). b, Correlation of cytoplasmic AR with total AR within pre-BAT biopsies (n=24). c, Correlation of AR mRNA with total AR within pre-BAT biopsies (n=15). d, Pre-BAT AR nuclear-to-cytoplasmic ratio (N:C) stratified by the presence or absence of a clinical response on C4D1 for samples adequate for IHC (n=24). Response defined as the presence of a PSA_50_ response or objective response on C4D1. p value by unpaired two-tailed t-test. For a-c, r and p values by Pearson’s correlation calculation.

**Extended Data Figure 3.**
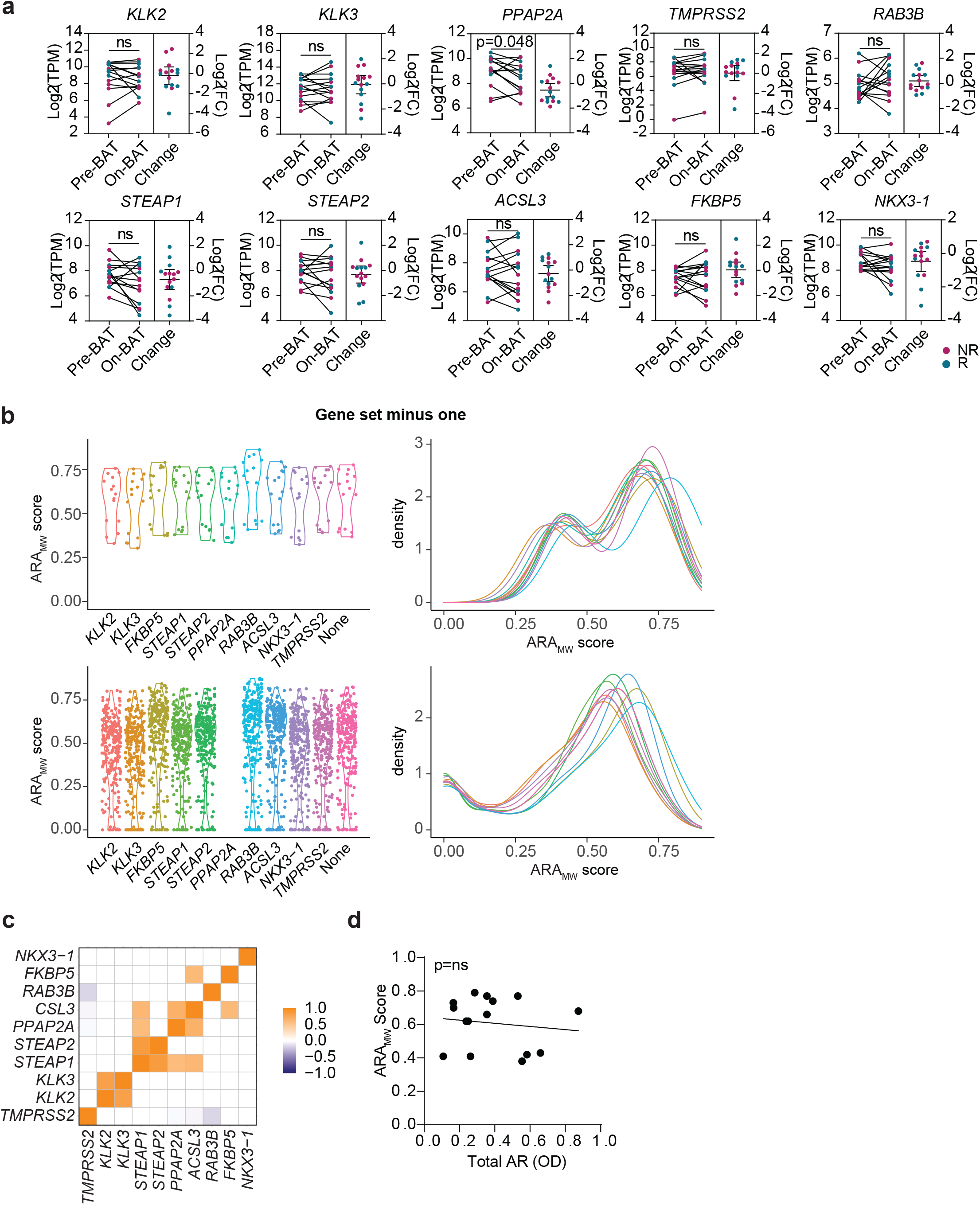
ARA_MW_ score reflects expression of a panel of AR target genes and does not correlate with AR mRNA expression. a, AR target gene mRNA expression by RNA sequencing of patient tumor biopsy samples pre-BAT and on C4D1 of BAT (n = 15). p value by paired two-tailed t-test. b, Distribution of ARA_MW_ scores in the COMBAT pre-BAT mCRPC (n=15) and SU2C/PCF mCRPC (n=266) cohorts subtracting one gene from the gene set for score calculation. c, Similarity matrix of gene expression of genes used in the ARA_MW_ score among patients in the COMBAT pre-BAT mCRPC cohort (n=15). Scale is Pearson correlation coefficient. d, Correlation of ARA_MW_ score with AR protein expression among patients in the COMBAT pre-BAT mCRPC cohort (n=15). p value by Pearson’s correlation calculation.

**Extended Data Figure 4.**
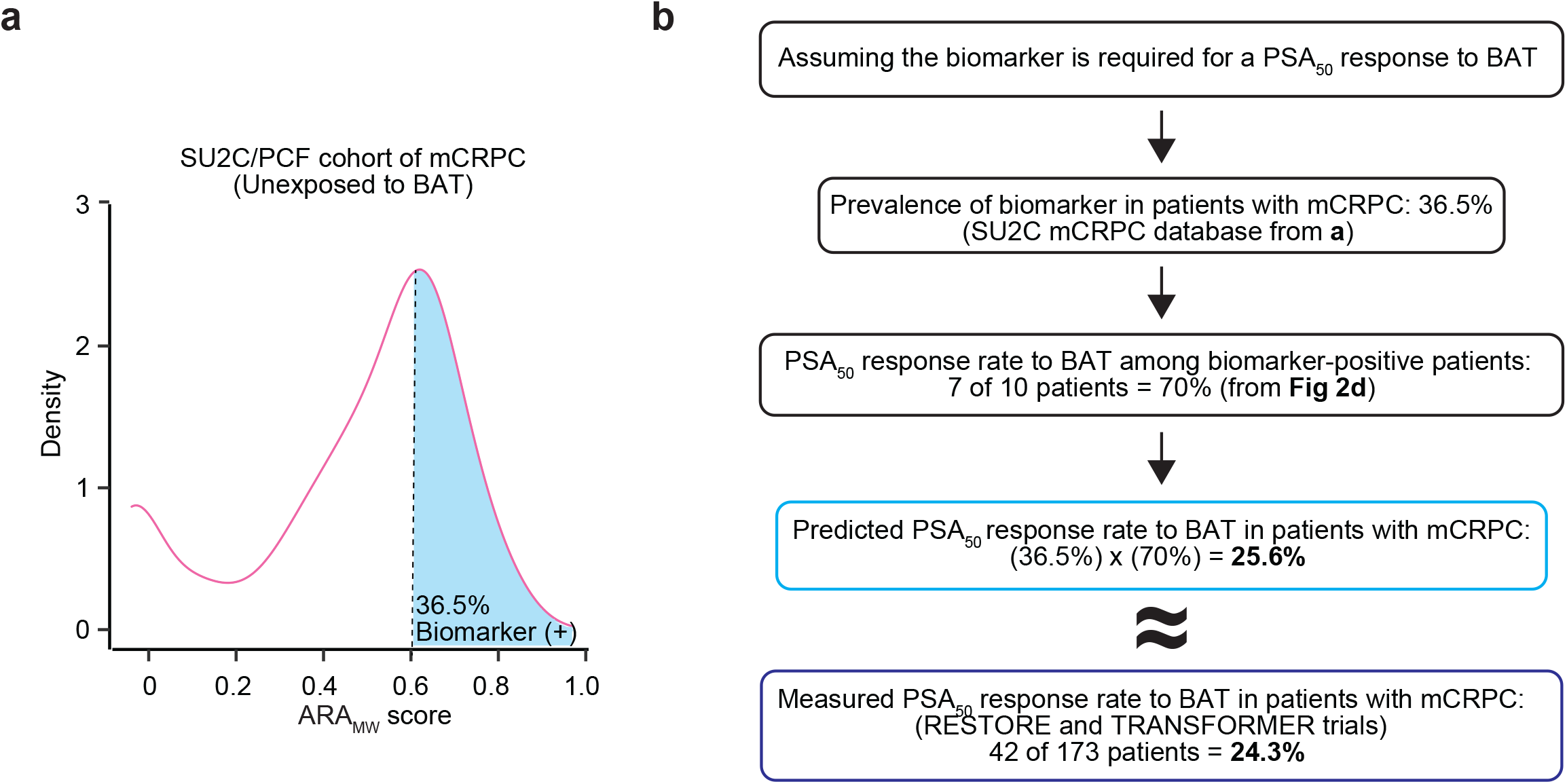
The prevalence of the biomarker accurately predicts efficacy of BAT. a, Distribution of ARA_MW_ score among patients in the SU2C/PCF cohort of mCRPC (n=266). Patients with a score greater than 0.6 assigned to be biomarker-positive. b, Flow chart to compare the estimated PSA_50_ response rate to BAT based on the prevalence of the biomarker versus the measured PSA_50_ response rate to BAT among patients with mCRPC.

**Extended Data Figure 5:**
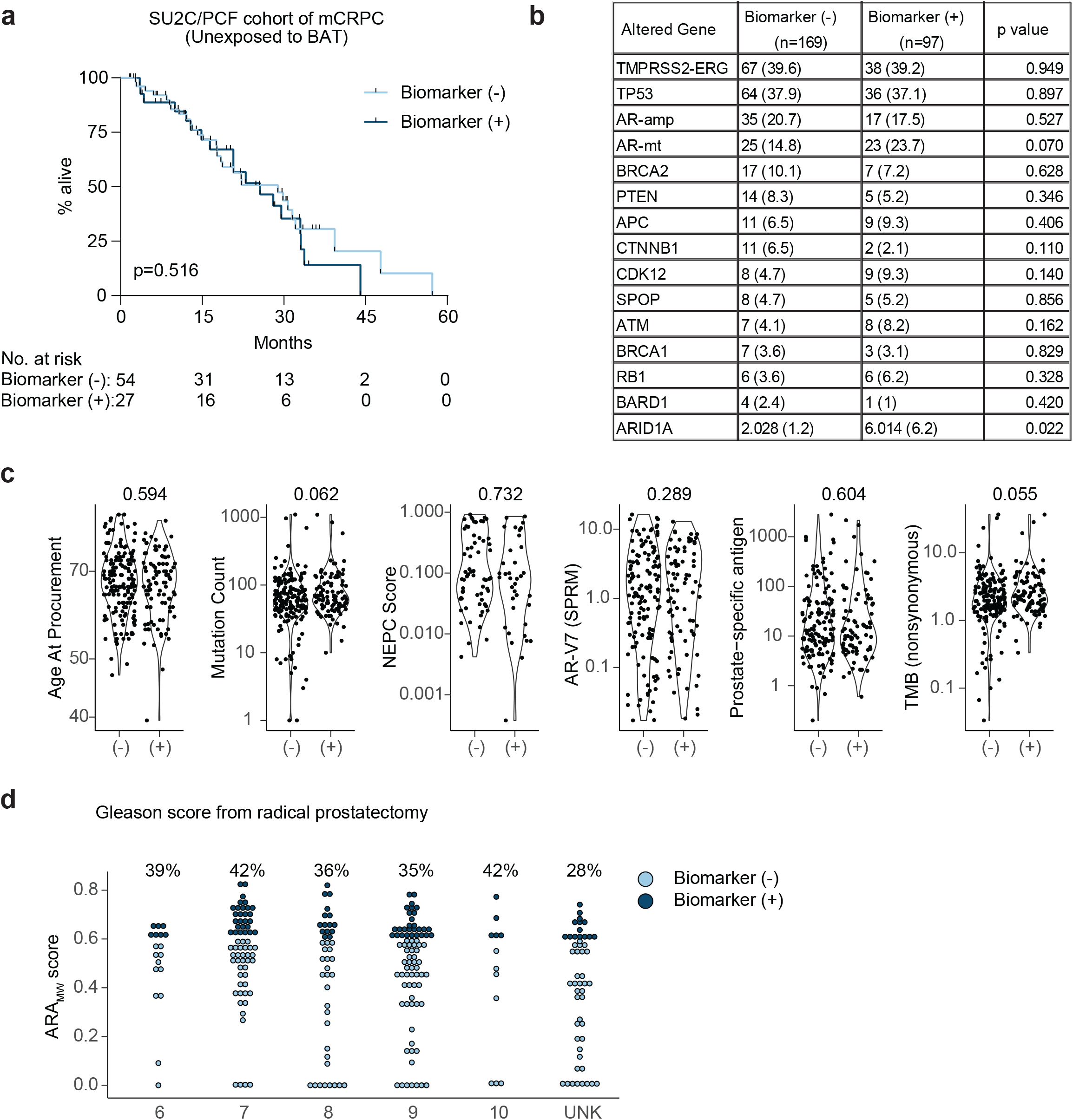
Biomarker-positive patients do not have inherently better prognosis in the absence of treatment with BAT. a, Overall survival of patients in the SU2C/PCF cohort stratified by presence or absence of the biomarker. p value by log-rank. b, Tumor gene mutations in patients in the SU2C/PCF cohort stratified by presence or absence of the biomarker. p value by Chi-squared comparison of proportions. c, Comparison of patient and tumor characteristics of patients in the SU2C/PCF cohort stratified by the presence or absence of the biomarker. p value by Wilcoxon ranked sum test. d, Gleason score of patients in the SU2C/PCF cohort stratified by the presence of absence of the biomarker. Biomarker (-), ARA_MW_ score less than 0.6. Biomarker (+), ARA_MW_ score greater than or equal to 0.6.

**Extended Data Figure 6.**
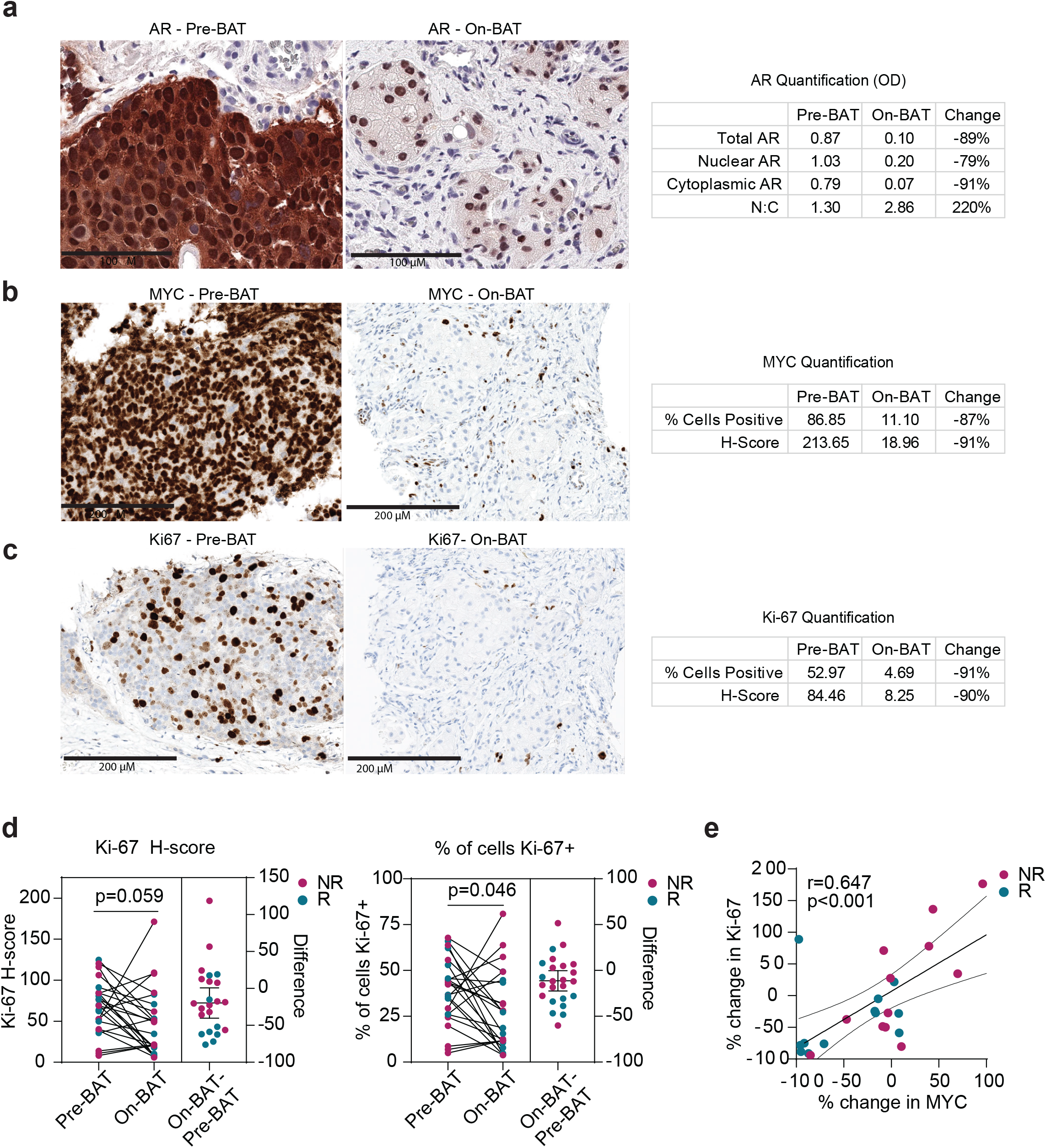
Change in MYC expression correlates with change in Ki-67 expression on BAT. Example of immunohistochemistry for AR (1:10,000 antibody dilution for quantitative image analysis) (a), MYC (b), and Ki-67 (c) in paired sequential biopsy samples from a responding patient. Quantification of protein in this patient’s samples are shown to the right of images. d, Ki-67 H-score and percentage of cells positive for Ki-67 in paired tumor biopsies suitable for IHC (n = 24 paired biopsies). p value by paired two-tailed t-test. e, Correlation of percent change in Ki-67 with percent change in MYC. Percent change in MYC truncated at 100%. r and p values by Pearson’s correlation calculation. NR, non-responder. R, responder.

**Extended Data Figure 7.**
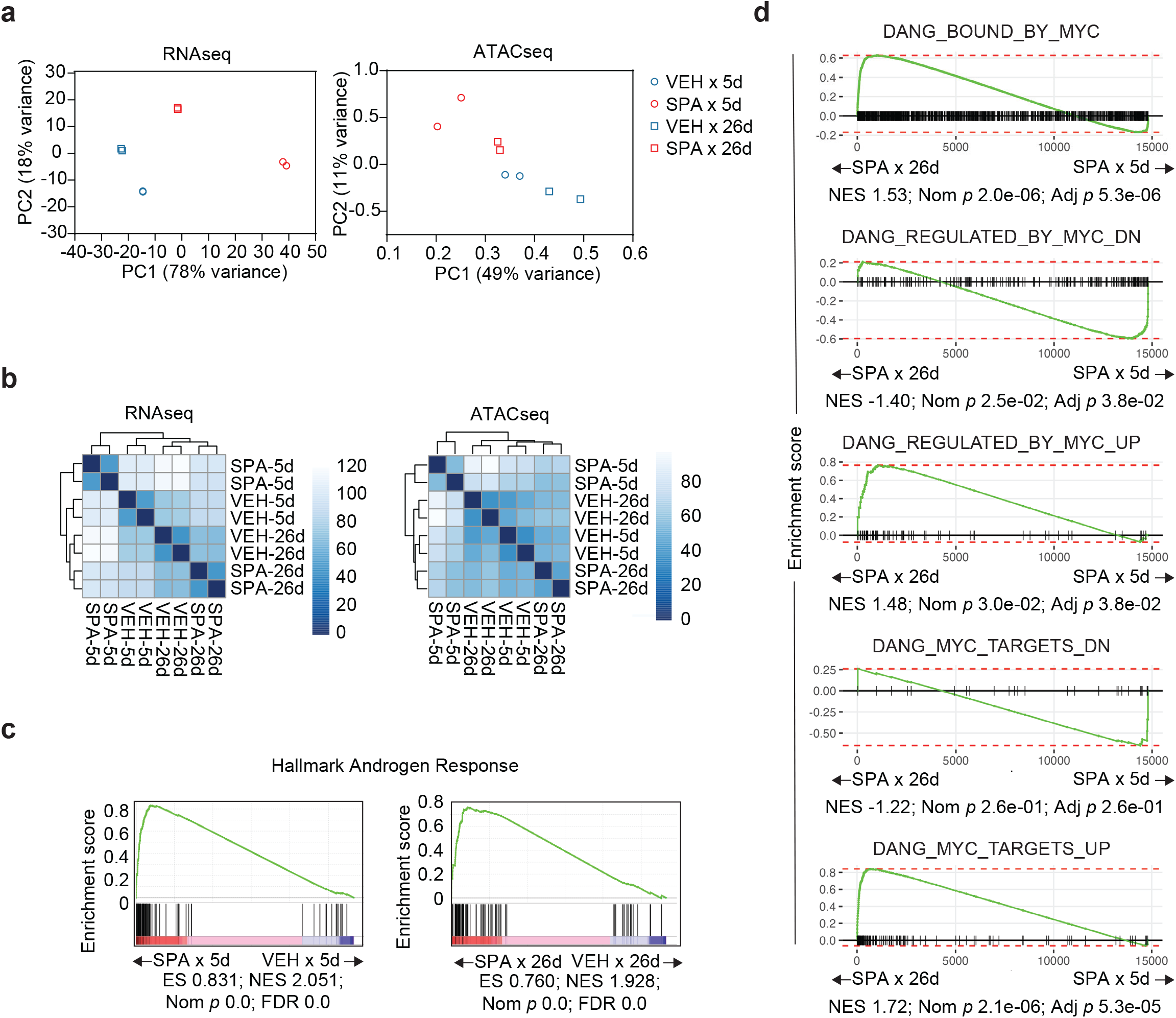
Cells with acquired resistance to SPA revert to a pre-treatment phenotype but maintain AR activation. Principal component (PC) analysis (a) and clustering analysis (b) of RNA sequencing and ATAC sequencing of LNCaP cells treated with VEH or SPA for 5 or 26 days. c, Hallmark Androgen Response gene set enrichment analysis tracings comparing LNCaP cells treated with VEH to SPA for 5 or 26 days. d, Gene set enrichment analysis tracings using gene sets previously determined to be regulated by MYC comparing LNCaP cells treated with SPA for 5 or 26 days. ES, enrichment score; NES normalized enrichment score; FDR, false discovery ratio.

**Extended Data Figure 8.**
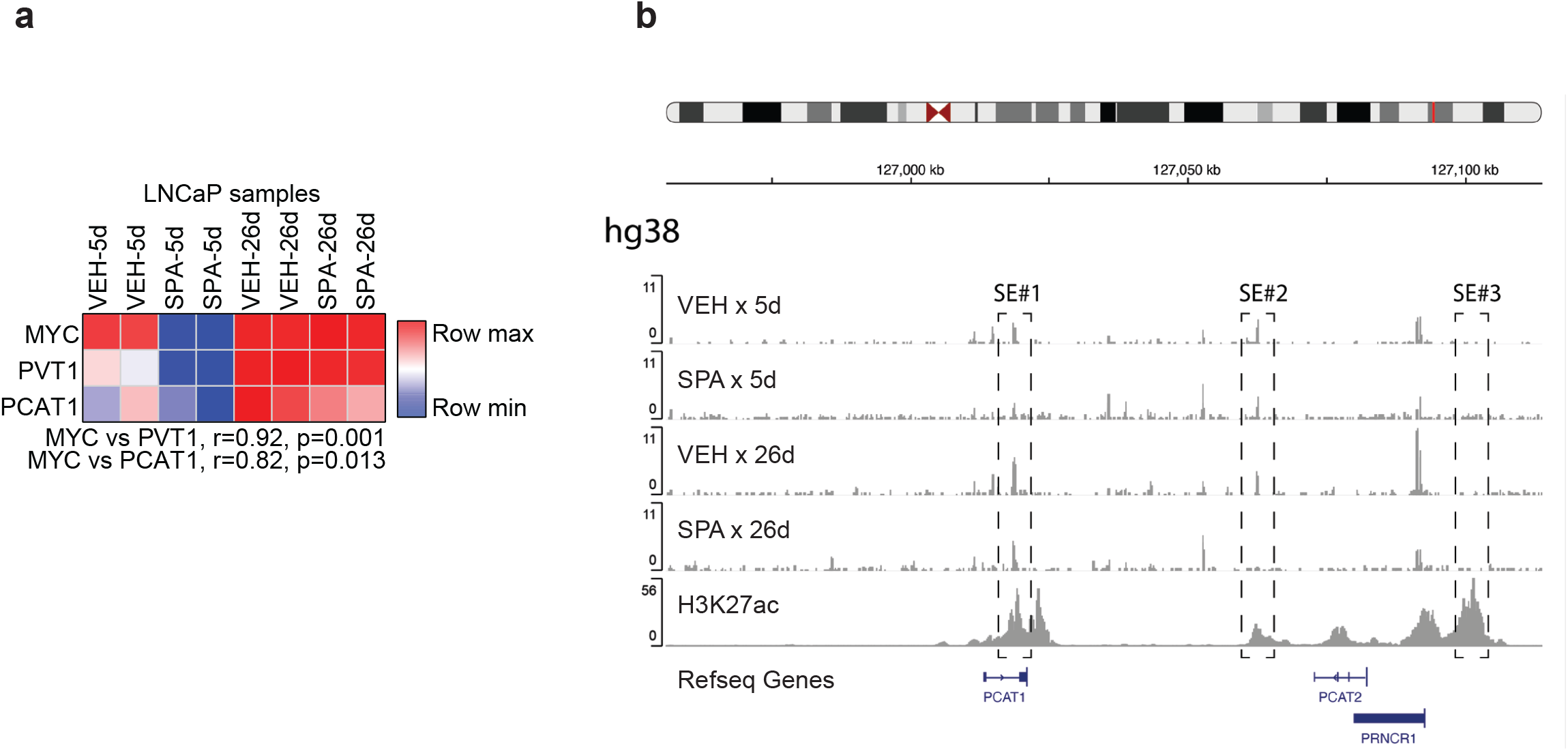
Acquired resistance to SPA is associated with an alteration in superenhancer activity on 8q24. a, Expression of genes within the 8q24 topologically associated domain (TAD) in LNCaP cells treated with VEH or SPA for 5 and 26 days. r and p values by Pearson’s correlation calculation. b, Chromatin accessibility of the MYC super-enhancers (SE) of LNCaP cells treated with VEH or SPA for 5 or 26 days. H3K27ac CHIP-seq on VCaP obtained from the ENCODE project. VEH, vehicle control, EtOH 0.01%. SPA, R1881 10 nM.

**Extended Data Figure 9.**
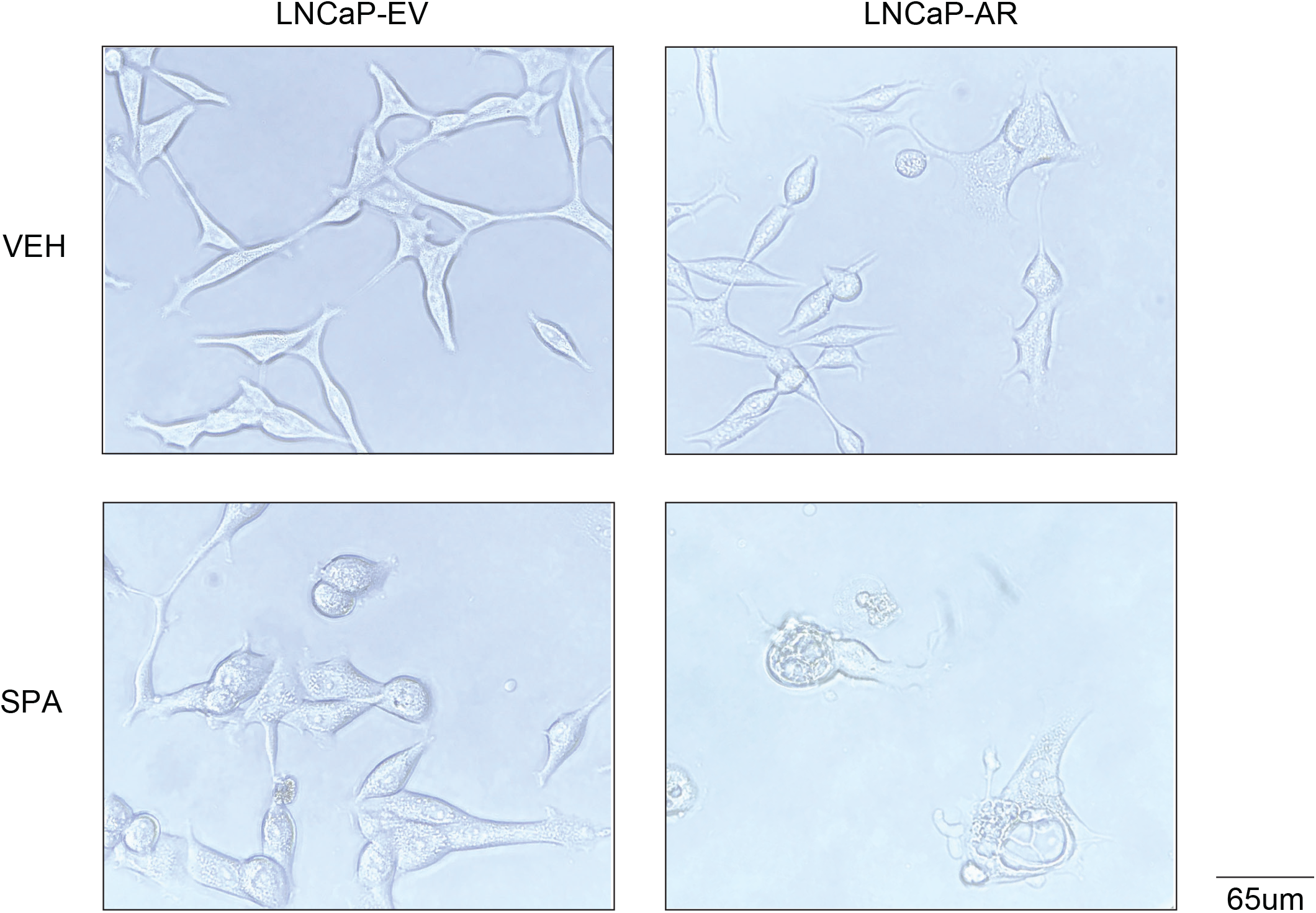
SPA results in extensive vacuolization in LNCaP cells with constitutively high AR expression. Representative photographs of light microscopy of LNCaP-EV and LNCaP-AR treated with VEH or SPA for 5 days. VEH, vehicle control, EtOH 0.01%. SPA, R1881 10 nM.

**Extended Data Figure 10.**
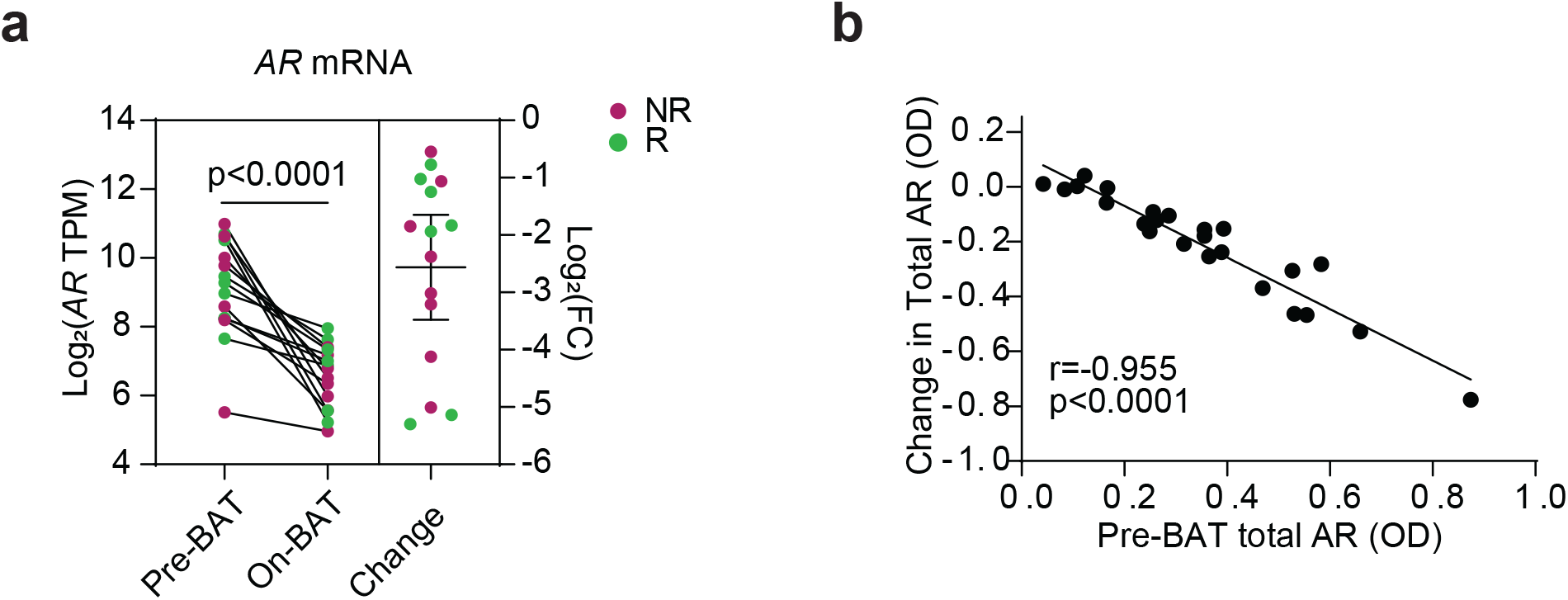
BAT downregulates AR expression. a, *AR* mRNA expression by RNA sequencing of patient tumor biopsy samples pre-BAT and on C4D1 of BAT (n = 15). p value by paired two-tailed t-test. Change with mean with 95% CI. b, Correlation of change in total AR optical density (OD) from pre-BAT to C4D1 of BAT to pre-BAT total AR OD (n=24). r and p values by Pearson’s correlation.

## References

[1] Schweizer MT, Antonarakis ES, Wang H, et al. Effect of bipolar androgen therapy for asymptomatic men with castration-resistant prostate cancer: results from a pilot clinical study. Sci Transl Med. 2015;7:269ra2.

[2] Teply BA, Wang H, Luber B, et al. Bipolar androgen therapy in men with metastatic castration-resistant prostate cancer after progression on enzalutamide: an open-label, phase 2, multicohort study. Lancet Oncol. 2018;19:76–86.

[3] Markowski MC, Wang H, Sullivan R, et al. A multicohort open-label phase II trial of bipolar androgen therapy in men with metastatic castration-resistant prostate cancer (RESTORE): A comparison of post-abiraterone versus post-enzalutamide cohorts. Eur Urol [Internet]. 2020; Available from: http://dx.doi.org/10.1016/j.eururo.2020.06.042.

[4] Sena LA, Wang H, Lim ScM SJ, et al. Bipolar androgen therapy sensitizes castration-resistant prostate cancer to subsequent androgen receptor ablative therapy. Eur J Cancer. 2020;144:302–309.

[5] Denmeade SR, Wang H, Agarwal N, et al. TRANSFORMER: A randomized phase II study comparing bipolar androgen therapy versus enzalutamide in asymptomatic men with castration-resistant metastatic prostate cancer. J Clin Oncol. 2021;JCO2002759.

[6] Kokontis J, Takakura K, Hay N, et al. Increased androgen receptor activity and altered c-myc expression in prostate cancer cells after long-term androgen deprivation. Cancer Res. 1994;54:1566–1573.

[7] Umekita Y, Hiipakka RA, Kokontis JM, et al. Human prostate tumor growth in athymic mice: inhibition by androgens and stimulation by finasteride. Proc Natl Acad Sci U S A. 1996;93:11802–11807.

[8] Denmeade SR, Isaacs JT. Bipolar androgen therapy: the rationale for rapid cycling of supraphysiologic androgen/ablation in men with castration resistant prostate cancer. Prostate. 2010;70:1600–1607.

[9] Sonnenschein C, Olea N, Pasanen ME, et al. Negative controls of cell proliferation: human prostate cancer cells and androgens. Cancer Res. 1989;49:3474–3481.

[10] Kumar R, Mendonca J, Owoyemi O, et al. Supraphysiologic testosterone induces ferroptosis and activates immune pathways through nucleophagy in prostate cancer. Cancer Res. 2021;81:5948–5962.

[11] Quarmby VE, Beckman WC Jr, Wilson EM, et al. Androgen regulation of c-myc messenger ribonucleic acid levels in rat ventral prostate. Mol Endocrinol. 1987;1:865–874.

[12] Ling MT, Chan KW, Choo CK. Androgen induces differentiation of a human papillomavirus 16 E6/E7 immortalized prostate epithelial cell line. J Endocrinol. 2001;170:287–296.

[13] Antony L, van der Schoor F, Dalrymple SL, et al. Androgen receptor (AR) suppresses normal human prostate epithelial cell proliferation via AR/β-catenin/TCF-4 complex inhibition of c-MYC transcription. Prostate. 2014;74:1118–1131.

[14] Wolf DA, Schulz P, Fittler F. Synthetic androgens suppress the transformed phenotype in the human prostate carcinoma cell line LNCaP. Br J Cancer. 1991;64:47–53.

[15] Wolf DA, Kohlhuber F, Schulz P, et al. Transcriptional down-regulation of c-myc in human prostate carcinoma cells by the synthetic androgen mibolerone. Br J Cancer. 1992;65:376–382.

[16] Lam H-M, Nguyen HM, Labrecque MP, et al. Durable response of enzalutamide-resistant prostate cancer to supraphysiological testosterone is associated with a multifaceted growth suppression and impaired DNA damage response transcriptomic program in patient-derived xenografts. Eur Urol. 2020;77:144–155.

[17] Guo H, Wu Y, Nouri M, et al. Androgen receptor and MYC equilibration centralizes on developmental super-enhancer. Nat Commun. 2021;12:7308.

[18] Gurel B, Iwata T, Koh CM, et al. Nuclear MYC protein overexpression is an early alteration in human prostate carcinogenesis. Mod Pathol. 2008;21:1156–1167.

[19] Koh CM, Gurel B, Sutcliffe S, et al. Alterations in nucleolar structure and gene expression programs in prostatic neoplasia are driven by the MYC oncogene. Am J Pathol. 2011;178:1824–1834.

[20] Litvinov IV, Vander Griend DJ, Antony L, et al. Androgen receptor as a licensing factor for DNA replication in androgen-sensitive prostate cancer cells. Proc Natl Acad Sci U S A. 2006;103:15085–15090.

[21] Haffner MC, Aryee MJ, Toubaji A, et al. Androgen-induced TOP2B-mediated double-strand breaks and prostate cancer gene rearrangements. Nat Genet. 2010;42:668–675.

[22] Chatterjee P, Schweizer MT, Lucas JM, et al. Supraphysiological androgens suppress prostate cancer growth through androgen receptor–mediated DNA damage. J Clin Invest. 2019;129:4245–4260.

[23] Robinson D, Van Allen EM, Wu Y-M, et al. Integrative Clinical Genomics of Advanced Prostate Cancer. Cell. 2015;162:454.

[24] Abida W, Cyrta J, Heller G, et al. Genomic correlates of clinical outcome in advanced prostate cancer. Proc Natl Acad Sci U S A. 2019;116:11428–11436.

[25] Cai C, He HH, Chen S, et al. Androgen receptor gene expression in prostate cancer is directly suppressed by the androgen receptor through recruitment of lysine-specific demethylase 1. Cancer Cell. 2011;20:457–471.

[26] Boumahdi S, de Sauvage FJ. The great escape: tumour cell plasticity in resistance to targeted therapy. Nat Rev Drug Discov. 2020;19:39–56.

[27] Zhu Y, Dalrymple SL, Coleman I, et al. Role of androgen receptor splice variant-7 (AR-V7) in prostate cancer resistance to 2nd-generation androgen receptor signaling inhibitors. Oncogene. 2020;39:6935–6949.

[28] Stanková K, Brown JS, Dalton WS, et al. Optimizing cancer treatment using game theory: A review. JAMA Oncol. 2019;5:96–103.

[29] Linja MJ, Savinainen KJ, Saramäki OR, et al. Amplification and overexpression of androgen receptor gene in hormone-refractory prostate cancer. Cancer Res. 2001;61:3550–3555.

[30] Scher HI, Sawyers CL. Biology of progressive, castration-resistant prostate cancer: directed therapies targeting the androgen-receptor signaling axis. J Clin Oncol. 2005;23:8253–8261.

[31] Markowski MC, Taplin M-E, Aggarwal RR, et al. COMBAT-CRPC: Concurrent administration of bipolar androgen therapy (BAT) and nivolumab in men with metastatic castration-resistant prostate cancer (mCRPC). J Clin Oncol. 2021;39:5014–5014.

[32] Gendusa R, Scalia CR, Buscone S, et al. Elution of high-affinity (>10-9 KD) antibodies from tissue sections: Clues to the molecular mechanism and use in sequential immunostaining. J Histochem Cytochem. 2014;62:519–531.

[33] Trabzonlu L, Kulac I, Zheng Q, et al. Molecular pathology of high-grade prostatic intraepithelial neoplasia: Challenges and opportunities. Cold Spring Harb Perspect Med [Internet]. 2019;9. Available from: http://dx.doi.org/10.1101/cshperspect.a030403.

[34] Freeman ZT, Nirschl TR, Hovelson DH, et al. A conserved intratumoral regulatory T cell signature identifies 4-1BB as a pan-cancer target. J Clin Invest. 2020;130:1405–1416.

[35] Li B, Dewey CN. RSEM: accurate transcript quantification from RNA-Seq data with or without a reference genome. BMC Bioinformatics. 2011;12:323.

[36] Dobin A, Davis CA, Schlesinger F, et al. STAR: ultrafast universal RNA-seq aligner. Bioinformatics. 2013;29:15–21.

[37] Ramírez F, Ryan DP, Grüning B, et al. deepTools2: a next generation web server for deep-sequencing data analysis. Nucleic Acids Res. 2016;44:W160–5.

[38] Liao Y, Smyth GK, Shi W. featureCounts: an efficient general purpose program for assigning sequence reads to genomic features. Bioinformatics. 2014;30:923–930.

[39] Love MI, Huber W, Anders S. Moderated estimation of fold change and dispersion for RNA-seq data with DESeq2. Genome Biol. 2014;15:550.

[40] Korotkevich G, Sukhov V, Budin N, et al. Fast gene set enrichment analysis [Internet]. bioRxiv. bioRxiv; 2016. Available from: http://dx.doi.org/10.1101/060012.

[41] Andreatta M, Carmona SJ. UCell: Robust and scalable single-cell gene signature scoring. Comput Struct Biotechnol J. 2021;19:3796–3798.

[42] Spratt DE, Alshalalfa M, Fishbane N, et al. Transcriptomic heterogeneity of androgen receptor activity defines a de novo low AR-active subclass in treatment naïve primary prostate cancer. Clin Cancer Res. 2019;25:6721–6730.

[43] Cerami E, Gao J, Dogrusoz U, et al. The cBio cancer genomics portal: an open platform for exploring multidimensional cancer genomics data. Cancer Discov. 2012;2:401–404.

[44] Bolger AM, Lohse M, Usadel B. Trimmomatic: a flexible trimmer for Illumina sequence data. Bioinformatics. 2014;30:2114–2120.

[45] Langmead B, Salzberg SL. Fast gapped-read alignment with Bowtie 2. Nat Methods. 2012;9:357–359.

[46] Li H, Handsaker B, Wysoker A, et al. The Sequence Alignment/Map format and SAMtools. Bioinformatics. 2009;25:2078–2079.

[47] Zhang Y, Liu T, Meyer CA, et al. Model-based analysis of ChIP-Seq (MACS). Genome Biol. 2008;9:R137.

[48] Ross-Innes CS, Stark R, Teschendorff AE, et al. Differential oestrogen receptor binding is associated with clinical outcome in breast cancer. Nature. 2012;481:389–393.

[49] Quinlan AR, Hall IM. BEDTools: a flexible suite of utilities for comparing genomic features. Bioinformatics. 2010;26:841–842.

[50] Robinson P, Zemojtel T. Integrative genomics viewer (IGV): Visualizing alignments and variants. Computational Exome and Genome Analysis. Chapman and Hall/CRC; 2017. p. 233–245.

[51] Zhang J, Lee D, Dhiman V, et al. An integrative ENCODE resource for cancer genomics. Nat Commun. 2020;11:3696.

[52] Zhu Y, Sharp A, Anderson CM, et al. Novel junction-specific and quantifiable in situ detection of AR-V7 and its clinical correlates in metastatic castration-resistant prostate cancer. Eur Urol. 2018;73:727–735.

[53] Raivo Kolde (2019). pheatmap: Pretty Heatmaps. R package version 1.0.12. https://CRAN.R-project.org/package=pheatmap

